# The clinical utility and epidemiological impact of self-testing for SARS-CoV-2 using antigen detecting diagnostics: a systematic review and meta-analysis

**DOI:** 10.1101/2022.07.03.22277183

**Authors:** Lukas E. Brümmer, Christian Erdmann, Hannah Tolle, Sean McGrath, Ioana D. Olaru, Stephan Katzenschlager, Seda Yerlikaya, Maurizio Grilli, Nira R. Pollock, Berra Erkosar, Aurelien Mace, Stefano Ongarello, Cheryl C. Johnson, Jilian A. Sacks, Claudia M. Denkinger

## Abstract

**Introduction:** Self-testing for COVID-19 (C19ST) based on antigen detecting diagnostics could significantly support controlling the SARS-CoV-2 pandemic. To inform the World Health Organization in developing a C19ST guideline, we performed a systematic review and meta-analysis of the available literature.

**Methods:** We electronically searched Medline and the Web of Science core collection, performed secondary reference screening, and contacted experts for further relevant publications. Any study published between December 1, 2020 and November 30, 2021 assessing the epidemiological impact and clinical utility of C19ST was included. Study quality was evaluated using the Newcastle Ottawa Scale (NOS). The review was registered on PROSPERO (CRD42022299977).

**Results:** 11 studies only from high-income countries with an overall low quality (median of 3/9 stars on the NOS) were found. Pooled C19ST positivity was 0.2% (95% CI 0.1% to 0.4%; eight data sets) in populations where otherwise no dedicated testing would have occurred. The impact of self-testing on virus transmission was uncertain. Positive test results mainly resulted in people having to isolate without further confirmation of results (eight data sets). When testing was voluntary by study design, pooled testing uptake was 53.2% (95% CI 36.7% to 68.9%; five data sets. Outside direct health impacts, C19ST reduced quarantine duration and absenteeism from work, and made study participants feel safer. Study participants favored self-testing and were confident that they performed testing and sampling correctly.

**Conclusions:** The present data suggests that C19ST could be a valuable tool in reducing the spread of COVID-19, as it can achieve good uptake, may identify additional cases, and was generally perceived as positive by study participants. However, data was very limited and heterogenous, and further research especially in low- and middle-income countries is needed to assess the clinical utility and epidemiological impact of C19ST in more detail.

**CONTRIBUTIONS TO THE LITERATURE:**

- COVID-19 self-testing (C19ST) using antigen detection could conceivably support pandemic control. A current PubMed search found no systematic evidence synthesis of studies assessing the epidemiological impact and clinical utility of C19ST implementation
- We systematically reviewed and meta-analyzed 11 studies including more than 1.1 million persons tested
- C19ST can achieve good uptake, may identify additional cases, and was general perceived as positive by study participants, suggesting it to be a valuable tool in reducing the spread of SARS-CoV-2
- Further data especially from low- and middle-income countries is needed to understand the impact of C19ST in more detail

## BACKGROUND

“We have a simple message for all countries: test, test, test.” [1] With this statement at the beginning of the COVID-19 pandemic, the World Health Organization’s (WHO) director general set the scene for what has developed into one of the main pillars for reducing the spread of SARS-CoV-2 [2]. Detecting cases early in the course of the disease and when pre- or asymptomatic has shown to be crucial for preventing further infections prior to the availability of vaccines [3-5]. Still, a large proportion of SARS-CoV-2 cases remain undiagnosed [6,7], or are only diagnosed after most secondary transmissions have already occurred [8].

For detecting SARS-CoV-2, antigen-based rapid diagnostic tests (Ag-RDTs), using lateral-flow assays, have become one of the main tools for testing. Compared to the gold-standard, real-time reverse transcriptase polymerase chain reaction (rRT-PCR), Ag-RDTs are less accurate in detecting SARS-CoV-2 infection, but still detect over 90% of the cases with high viral load in the early phase of disease [9]. In addition, they require minimal implementation resources and are easy to use [10], allowing their widespread application. On a population level, modelling studies show that the Ag-RDTs’ disadvantage of lower accuracy is outweighed by their potential to be implemented frequently and at large scale, thus conceivably detecting cases earlier, which could result in reducing the spread of SARS-CoV-2 [11,12]. Moreover, studies have shown Ag-RDTs to be similarly accurate when performed by lay users or medically-trained persons [13]. Consequently, Ag-RDTs have already been applied in multiple settings as a tool for COVID-19 self-testing (C19ST) [14-17]. By increasing available testing capacity, C19ST using antigen-detection diagnostics could be an additional tool for pandemic control if implemented at scale [18,19].

However, the clinical utility, i.e., the extent to which it improves health benefits, and the epidemiological impact of implementing C19ST using Ag-RDTs has not yet been systematically evaluated [20]. Aiming to fill this gap in the literature and to assist the WHO in developing a guideline for Ag-RDT based C19ST in March 2022 [19], we performed a systematic review of the existing literature.

## METHODS

A study protocol was developed (S1 Text), which follows standard guidelines for systematic reviews [21,22] and was reviewed by an independent methodologist contracted by the WHO. We completed the PRISMA checklist (S1 PRISMA Checklist) and registered the review on PROSPERO (registration number: CRD42022299977). This review was designed in close cooperation with the WHO and the members of the guideline development group [19].

### Search strategy

We searched for studies assessing the clinical utility and epidemiological impact of implementing self-performed Ag-RDTs for SARS-CoV-2. A professional librarian (MG) electronically searched MEDLINE (via PubMed) and Web of Science for any article published between December 1, 2020 and November 30, 2021. The main search terms were “Severe Acute Respiratory Syndrome Coronavirus 2”, “COVID-19”, “Betacoronavirus”, “Coronavirus”, “Self-testing”, “home test”, and “Antigen” (details can be found in the Supplements, S2 Text). No geographic or language restrictions were used.

### Study selection

Two reviewers (LEB and HT) independently reviewed the titles and abstracts of all publications identified through the electronic search. Afterwards, three reviewers (LEB, HT and CE) individually conducted a full-text review, to select the articles for inclusion in the systematic review. As a final step, the three reviewers compared results with each other (for all steps described using EndNote, version X9 [23]). Any disputes were resolved by discussion or by a senior reviewer (CMD).

To be included in the review, a study had to implement C19ST using Ag-RDTs in a symptomatic or asymptomatic population during the COVID-19 pandemic, compare this to a situation when no testing, professional use Ag-RDT testing or PCR-testing is performed, and measure one or more of the outcomes presented in Table 1, which were prioritized based on a survey with the prospective members of the WHO Guideline Development Group (a detailed explanation for each outcome is presented in the Supplement, S3 Text). We considered retrospective or prospective cohort or nested cohort studies, case-control or cross-sectional studies, before and after studies, as well as randomized studies. Where values and preferences for C19ST were reported by studies identified through the search, they were also descriptively summarized and reported (i.e., studies that assessed values and preferences only but without actually implementing C19ST were not considered). Studies with less than 100 study participants were excluded. The review was limited to studies among human participants.

**Table 1.** Outcomes as ranked by the members of the WHO guideline development group.

| Outcomes | Sub-outcomes | Score | Rank |
| --- | --- | --- | --- |
| (1) Community level health impact | a) Proportion of infectious cases detected / positive test results | 6 | Important |
|  | b) Impact on virus transmission | 6 | Important |
|  | c) Impact on morbidity and/or mortality | 6 | Important |
| (2) Individual level health impact | a) Linkage for positive tests / actions after positive test result | 7 | Critical |
|  | b) Social harm | 7 | Critical |
|  | c) Testing uptake | 6.5 | Critical |
|  | d) Time to diagnosis | 6.5 | Critical |
|  | e) Testing frequency | 6 | Important |
|  | f) Result reporting | 6 | Important |
|  | g) Behavior change | 5.5 | Important |
|  | h) Linkage for negatives / actions after negative test result | 5 | Important |
| (3) Broader societal effects |  | 6 | Important |

In addition, references of the studies included into the review were manually searched by two reviewers (LEB and HT) to identify further studies for inclusion. Furthermore, selected experts in the field were contacted to identify additional articles not identified through the electronic search. While the electronic search was limited to peer-reviewed data bases, we also considered any pre-print identified through secondary reference searching or contacting selected experts for inclusion.

### Data extraction and management

Data extraction was performed by one reviewer (HT), using a standardized Google Sheets form, and all extracted data was checked by a second reviewer (LEB). Any disagreements about the data extracted were resolved through consensus or referral to a senior study team member (CMD). Study authors were contacted if data was missing or there were questions on the reporting of the outcomes. Several data sets were extracted from studies that assessed multiple self-performed Ag-RDTs or presented results based on differing settings (e.g., school vs. workplace). A full list of variables extracted can be found in the Supplements (S1 Table).

### Quality assessment

To assess the quality of the included studies, the Newcastle-Ottawa Scale (NOS) was employed [24]. Accounting for intra-study heterogeneity, if several data sets were extracted from one study, these were assessed individually. Depending on the study’s quality, stars are assigned to each of the NOS’s questions, leading to an overall zero-star-rating (worst) to nine-star-rating (best) for each study. We prepared an interpretation guide specific for the research question of this review for each of the questions on the NOS, which can be found in the Supplements (S4 Text). The quality assessment was performed by two reviewers (LEB and HT) independently. Any disputes were solved by discussion or by a third reviewer (CMD).

### Data analysis

For quantitative outcomes, if four or more data sets were available, we prepared forest plots, visually evaluated the heterogeneity between studies and conducted a meta-analysis using random effects analysis. Specifically, we fit a random intercept logistic regression model to meta-analyze the proportion of infectious cases detected and mean testing uptake [25]. For outcomes where limited data was available, we performed a descriptive analysis.

We pre-defined the following categories to stratify outcomes and intended to meta-analyze based on these subgroup categories if reported by at least four papers: (1) frequency of testing (one-off testing vs testing more than once; for testing more than once: testing at random, i.e. without a fixed schedule, vs. testing as per a fixed schedule); (2) SARS-CoV-2 exposure (no known exposure, known exposure – single or multiple, unknown / not defined); (3) test distributor (workplace, school/university, publicly available); (4) location of testing (work, home, school/university); (5) cost per person to be tested (tests free of charge vs. payment required), (6) setting (rural or urban, gross domestic product (GDP) per capita [categorized by World Trade Organization], literacy of the target population), (7) Ag-RDT used (company name, test and lot number, sample type), (8) assistance in sample collection or performing the test (no assistance vs. assisted in person by trained operator vs. assisted virtually by trained operator vs. assisted in any other way), (9) symptomatic or asymptomatic study participants (10) risk of COVID-19 related morbidity and mortality (as defined by study), (11) vaccination status (vaccinated vs. unvaccinated), (12) prior infection (convalescent vs. no known infection), (13) age (persons ≤ 18 years of age, persons 18 – 65 years of age, persons > 65 years of age); (14) misuse, adverse events, social harms (specific to self-testing vs. issues that occur in general for SARS-CoV-2 testing), (15) additional interventions (any other interventions that were implemented to stop transmission besides C19ST for SARS-CoV-2, such as mask wearing or travel restrictions).

The analyses were performed in R (version 4.1.3) with package “meta”. The R code and the raw data are available at https://github.com/stmcg/covid-testing-impact-ma.

### Grading of evidence

Following the GRADE approach [26], we prepared an interpretation guide specific for the research question of this review for each of the GRADE aspects (S5 Text). Grading of evidence was performed by one reviewer (LEB) and reviewed by a second (CCJ). Any disputes were resolved by discussion or by a third reviewer (CMD). Using GRADE evidence profiles, a summary of the GRADE assessment for each outcome is presented in the results section, and a detailed assessment in the Supplements (S6 Text).

## RESULTS

### Study description

The search yielded 2,722 records. After removing duplicates, 1,826 unique records remained, of which 1,642 were excluded based on title and abstract screening. The full text of 182 articles was read, with seven being found to be eligible for inclusion in the review. Experts identified four additional reports, which were also included in the review. Overall, 11 studies were included [15-17,27-34], incorporating a total of 17 datasets (Fig 1). A list of the excluded studies can be found in the Supplement (S7 Text).

**Fig 1.**
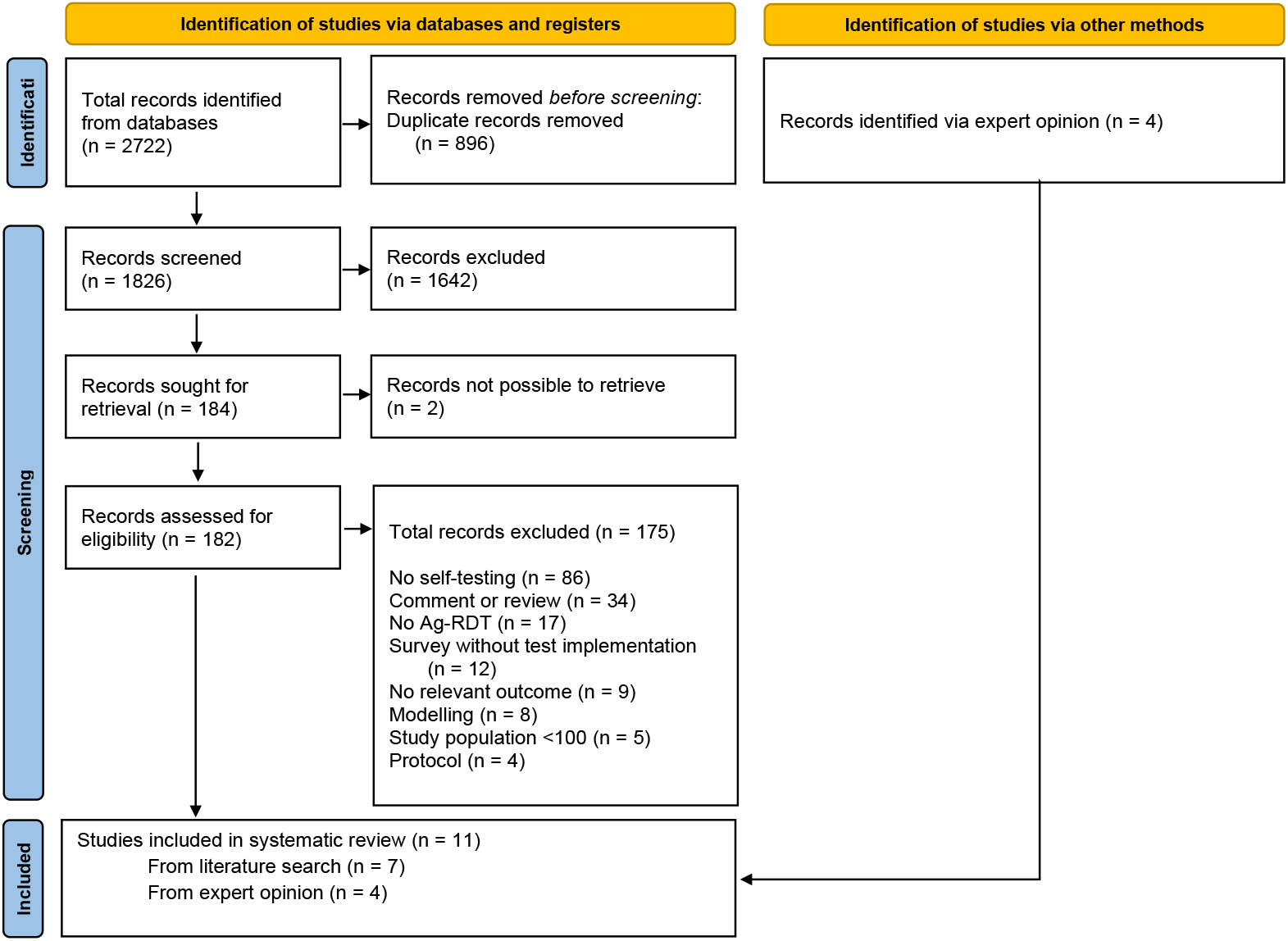
PRISMA flow diagram. Ag-RDT = antigen rapid diagnostic test

Most of the datasets were from the United Kingdom (8; 47.1% of all data sets) [15,16,28,29,33,34], and the others from Germany (4; 23.5% of all data sets) [17,27,32], Austria (3; 17.6% of all data sets) [31], and the Netherlands (2; 11.8% of all data sets) [30]. In eight data sets (47.1% of all data sets) [15,16,28,29,33,34], the Innova SARS-CoV-2 Rapid Antigen Self-Test Kit (Innova Medical Group Inc., United States [CA]; henceforth called ‘Innova’) was used. The STANDARD Q COVID-19 Ag Home Test (SD Biosensor, South Korea; distributed in Europe by Roche, Germany; henceforth called ‘Standard Q’) [30,35] and the SARS-CoV-2 Antigen Rapid Test Kit (Lepu Medical Technology, China) [31] were used in three data sets each (17.6% of all data sets). Of the remaining three data sets, one (5.9% of all data sets) used the BD Veritor At-Home COVID-19 Test (Becton, Dickinson and Company, United States [NJ]) [30], another the RIDA QUICK SARS-CoV-2 Antigen (R-Biopharm, German) [32] and for the third the Ag-RDT used was unclear [27]. In eleven data sets (64.7% of all data sets), the sample type was anterior nasal / mid-turbinate (AN/MT) [28-33,35], while the sample type used in the remaining six data sets (35.3% of all data sets) was not reported [15,16,27,34].

In 14 datasets (77.8% of all datasets) [15-17,27-29,31-34], C19ST was implemented as part of a routinely testing service, most often (4 datasets) [29,31,33,34] for testing twice weekly. C19ST was mainly performed at home (10 data sets; 58.8%), but also in school (4; 23.5% of all data sets) [27,31] or at work (1; 5.9% of all data sets) [34]. In two data sets (11.7% of all data sets) from the same study, testing was performed at dedicated testing sites at the beginning of the study and at home towards end of the study [16]. Study participation and self-testing was free-of-charge in five data sets (27.8% of all data sets) [28,31], and there was no mention of cost of tests in the other data sets.

From a total of 1,124,911 study participants, the majority (785,472; 69.8% of all participants) were school students and teachers, 325,655 (28.9%) were unselected study participants, i.e., from the general population, 13,050 (1.2%) were hospital and care home staff and 734 (0.1%) were university students and staff. For two data sets [15,27], the number of study participants was unclear. 2,343 (0.3%) of all study participants were reported to be symptomatic, while 794,417 (70.6%) were asymptomatic and for the remaining 328,151 (29.2%) the symptom status was unclear. Detailed characteristics of each data set included in the review are presented in the Supplement (S2 Table, sheets ‘study characteristics’ and ‘testing characteristics’).

### Quality assessment

Overall, studies achieved a median of three (IQR 2.0 to 3.0) out of nine stars on the Newcastle Ottawa scale, indicating an overall low study quality (Supplement, S1 Fig). As outlined above, studies were assessed in regards to the selection of their study population, the comparability between study populations, and the assessment of the outcomes of interest for this review. A detailed quality assessment for each study is presented in the Supplements (S1 Fig and S3 Table, sheet ‘study assessment’).

In two out of 17 datasets (11.8%) [30,34], the study population for C19ST was likely representative of the general population as it was sampled from the general population, enrolled consecutively and with study participation rates above 90%. The remaining 15 (88.2%) datasets focused on sub-groups of the population (e.g., students) [15-17,27-29,31-33]. Where studies included a comparator cohort, i.e., a cohort that did not perform C19ST (6 datasets, 35.3%) [27,28,30,33,34], this was always drawn from the same population as the exposed cohort. The C19ST result was directly confirmed by study personnel (through observation of the testing procedure) or review of an upload a picture of the test result in three datasets (17.6%) [15,16]. For the remaining 14 (82.3%) datasets, test results were not confirmed (12 data sets) [15-17,28-32] or the process of documenting results was not further described (2 data sets) [27,34]. In 12 datasets (70.6%) [15-17,28,29,31,32], no other dedicated testing alternative were offered to participants (i.e., counterfactual was no testing at all) with initiation of the study. However, in five datasets (29.4%) [27,30,33,34], studies were conducted in settings where persons were also tested by RT-PCR.

Two datasets (11.8%) excluded persons who had a positive SARS-CoV-2 test result within the previous two months [27,29], enhancing comparability between study populations. In all other datasets, persons were included irrespective of prior infection or their vaccination status.

In three data sets (17.6%) [15,16], study personnel collected the data relevant to report on the outcomes. In the remaining 14 datasets (82.4%) relevant data was only self-reported by study participants (12 datasets) [15,17,28-33] or the procedure of data collection was unclear (2 data sets) [27,34]. In all but one dataset (94.1%) [33] the study period was clearly defined, suggesting that follow up was long enough for outcomes to occur. In three datasets (17.6%) [16,17,27], study participants were followed up adequately, i.e., for routine testing >80% of study participants followed the intended testing frequency across the whole study period. In four further data sets (23.5%) the loss to follow up was only minimal and therefor unlikely to introduce bias [15,31]. However, in the remaining 10 datasets (58.8%) [28-30,32-34] study results might have been biased due to loss of participants over the course of the study.

### Community level health impact

#### Proportion of cases detected / test positivity

The number and proportion of positive C19ST results was reported in eight datasets. All were in settings where no dedicated testing was performed before. The meta-analyzed proportion of positive test results was 0.2%(95% CI 0.1% to 0.4%) [15,16,29,31,32,34]. Test positivity ranged from 0.7% in the general population of a large city in the UK (Liverpool) [15] to 0.0% in university-wide screening, also in the UK [16] (Fig 2). The data was overall limited and from only observational studies with low quality (median of 3 stars on the NOS; IQR 2.8 to 3.3), resulting in a very low certainty of evidence (Supplements S6 Text).

**Fig 2.**
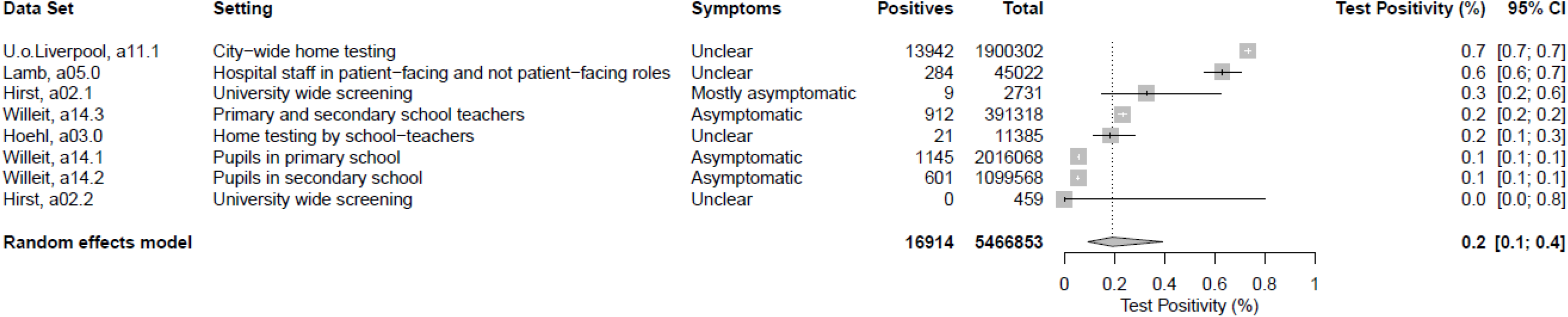
Test positivity in settings where no testing was performed before.

#### Impact on virus transmission

In one data set rRT-PCR was used to test care-home staff and inhabitants weekly or monthly, respectively, or if symptomatic. In addition, in selected care-homes (henceforth called ‘study care-homes’) C19ST was offered to care-home staff twice weekly. Also, visitors who tested negative through professional testing were allowed indoor visits. An increased proportion of outbreaks, defined as at least two confirmed or clinically suspected SARS-CoV-2 cases within two weeks, occurred in the study care homes (6 out of the 11 study care homes, i.e., 54.5% [95% CI 23.4% to 83.3%]) compared to the standard care homes that did not introduce self-testing for staff or allow indoor visits (26 out of 71 standard care homes, i.e., 36.6% [95% CI 25.5% to 48.9%]). However, confidence intervals were widely overlapping (Fig 3a) and it was not possible to distinguish whether differences were due to implementing C19ST for care-home staff or allowing indoor visits. Of note, in one of the six study care-homes experiencing an outbreak this was identified through C19ST, while in the other five the index case was identified through routine or symptomatic rRT-PCR testing [34].

**Fig 3a and 3b.**
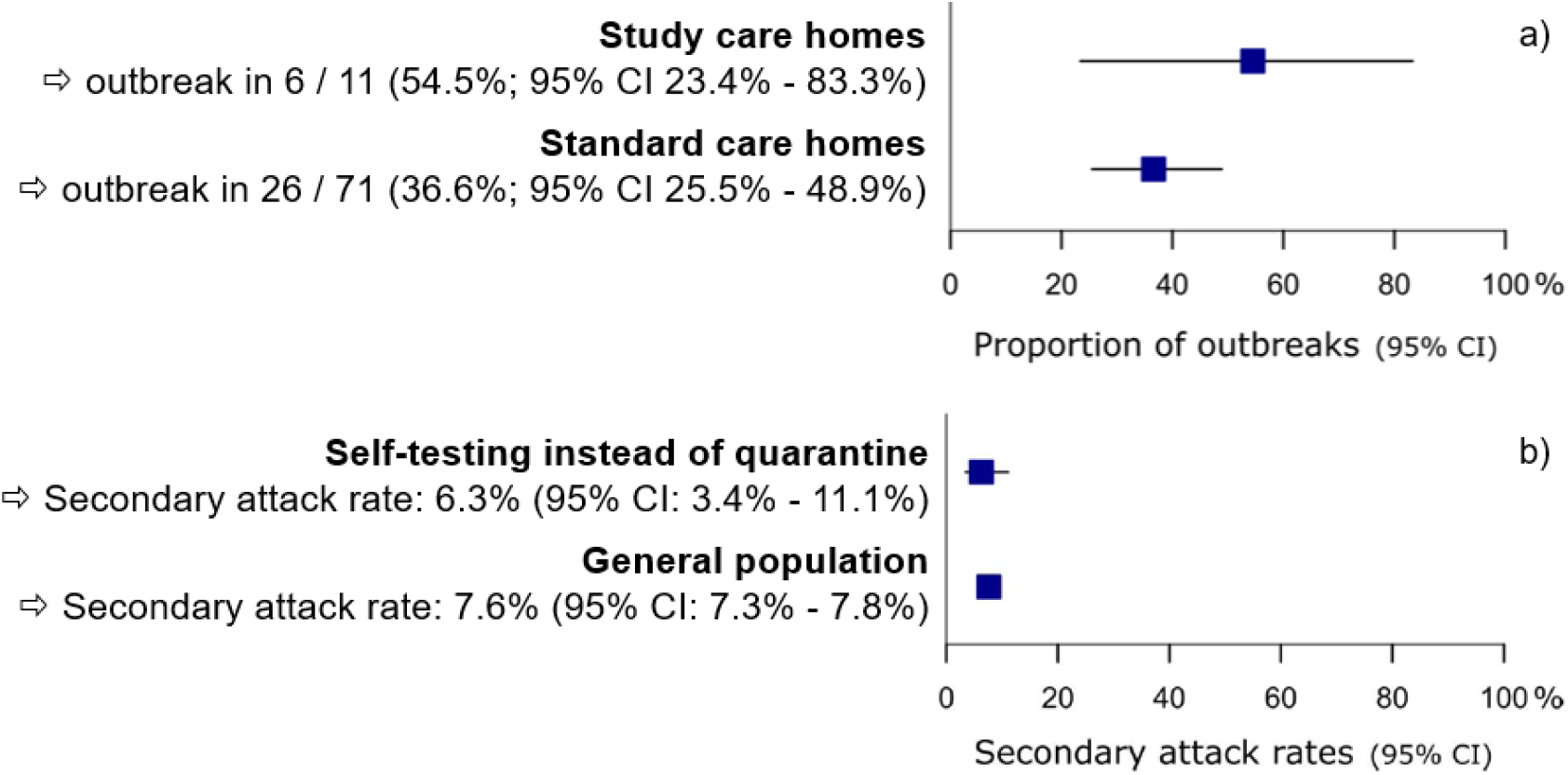
C19ST impact on virus transmission. CI = confidence intervals.

In a separate study, adults who had been exposed to a confirmed case were offered daily self-testing as an alternative to quarantine. Secondary attack rates were 6.3% (95% CI 3.4% to 11.1%) when Ag-RDTs were used instead of quarantine and positive results were confirmed by RT-PCR. In the comparator group, where quarantine was followed by a RT-PCR at day 7, the secondary attack rate in contacts of study participants was similar 7.6% (95% CI 7.3% to 7.8%) [28] (Fig 3b). Overall, data was too limited to establish an association between self-testing and virus transmission when implemented in a care-home or offered as an alternative to quarantine after contact with cases (Supplement S6 Text).

### Individual level health impact

#### Linkage for positive tests

Actions after a positive test result were reported in 11 datasets across two different settings. In nine data sets, positive results in mostly asymptomatic people required isolation (eight data sets) [16,17,29,31] or an invitation to seek confirmatory rRT-PCR testing (one data set) [15]. Second, in two datasets, positive test results of contacts already in quarantine required isolation [15,28]. In a total of five datasets [15,16,28,29], persons were required to seek a confirmatory rRT-PCR test after a positive C19ST result. Where results of the confirmatory rRT-PCR testing were reported, this is listed in the Supplements (S2 Table, sheet ‘testing characteristics’). Based on the limited data available, it is uncertain if self-testing affected actions after a positive test result in a different way compared to professional testing (Supplements S6 Text).

#### Testing uptake

In five datasets (10,336 individuals) testing was voluntary and the health impact of self-testing in the community was examined [16,17,28,29]. Across these datasets, testing uptake was estimated to be 53.2% (95% CI 36.7% to 68.9%), ranging from 75.7% in hospital staff [29] to 25.8% in university students and staff [16] (Fig 4). However, all five datasets were from observational studies with a low quality (median of 3 stars on the NOS; IQR 3 to 3). Thus, evidence was judged to be of very low certainty (Supplements S6 Text).

**Fig 4.**
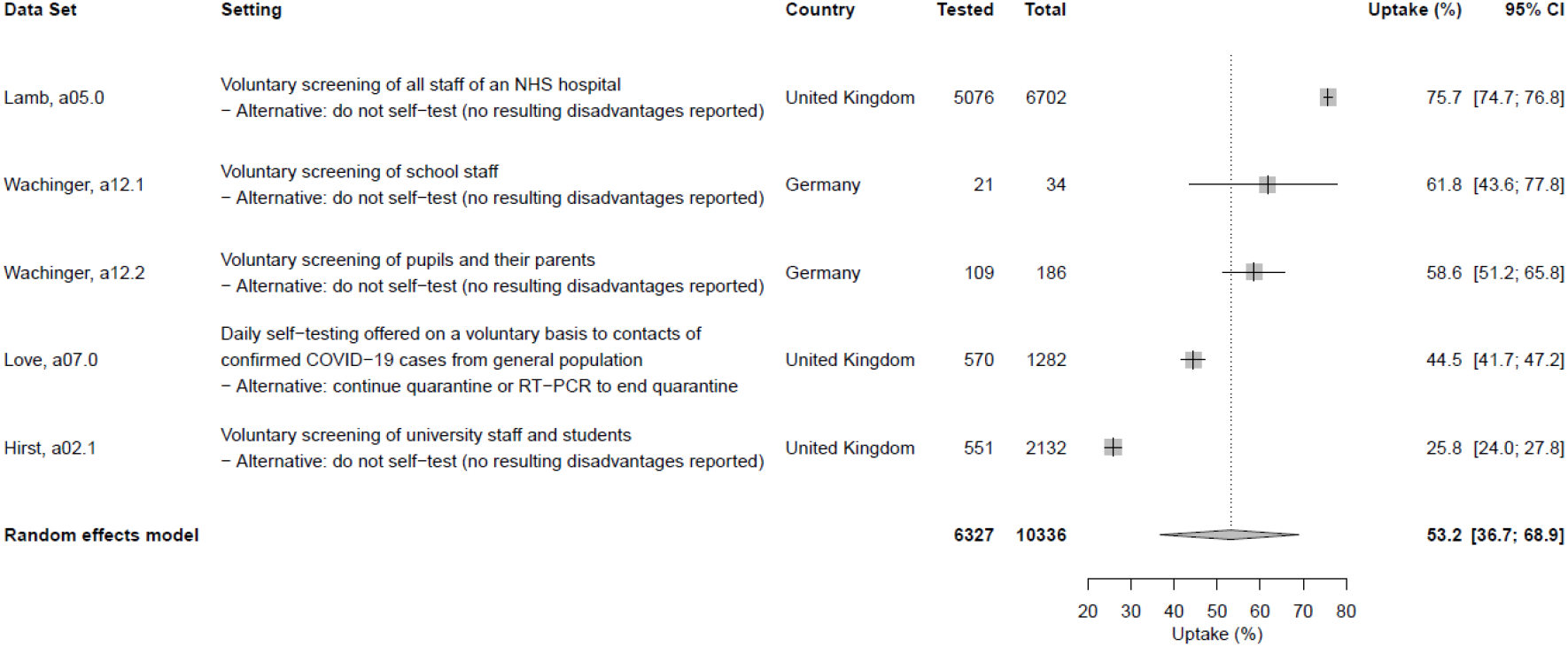
Testing uptake with self-test offer.

#### Time to diagnosis

In eight data sets, positive C19ST results were used to inform the need for isolation. In three of these, self-testing was implemented for one-off testing in hospital staff and university students and staff [16,29], while in the other five it was implemented for regular screening in schools [17,31]. With self-testing providing results within 20-30 minutes [36,37], the time to inform the need for isolation can be assumed to fall in a similar range. However, self-testing was not reported to be considered a final SARS-CoV-2 diagnosis, e.g., for clinical decision making or official documentation, in any of the datasets. As the data come from only observational studies with low quality (median of 3 stars on the NOS; IQR 3 to 3.2), the evidence to support this was of very low certainty (Supplements S6 Text).

#### Result reporting

Reporting the test result following self-testing was required by study design in four data sets [27,31]. Participants included in two datasets were contacted by telephone if they did not submit a test result and the proportion of results reported was 90.7% [30]. The proportion of results reported was unclear in 10 data sets [15-17,28,29,32-34]. It was not possible to differentiate between the proportion of results reported according positive or negative self-testing result. Again, overall, there was insufficient evidence to estimate the proportion of results reported in the general population or subgroups, but in the available data the proportion reported was high (> 90%) (Supplements S6 Text).

#### Linkage for negatives

Negative C19ST results were followed by the continuation of status-quo (e.g., continue with school operation, continue to work; 8 datasets) [16,17,31,33] or the permission to leave quarantine for 24 hours (2 datasets) [15,28]. In eight of these datasets, people had to retest within a given time interval (range 1 to 7 days) [15,17,28,31,33]. Based on the limited data available, it is uncertain if self-testing affects actions after a negative test result in a different way compared to professional testing (Supplements S6 Text).

### Broader societal effects

C19ST was reported to be linked to reduced quarantine duration (2 datasets) [15,28] and, consequently, decreased absenteeism from work (1 dataset) [15]. Self-testing also enabled primary school students and staff to feel safer when having in-person lessons (2 data sets) [17]. It is uncertain if self-testing affects absenteeism and provides other social benefits compared to professional testing alone due to very low certainty evidence (Supplements S6 Text). No reports of social harm or adverse events following C19ST were identified in the included data sets.

### Further outcomes

Additional data was not available for multiple outcomes, including impact on morbidity and mortality. Details are presented in the Supplements (S8 Text).

### Values and preferences

Three of the studies included in this review also reported on values and preferences towards self-testing [28,30,34]. Furthermore, we found two studies, which did not assess any of our review’s outcomes [38,39], but reported the values and preferences of the study participants from two of the studies that were included in our review [16,28].

When asking study participants from the general population, who were offered daily self-testing with AN/MT swabs as an alternative to quarantine after contact with a case [28], almost 70% had no concerns about performing daily self-testing [38]. In another study, persons also from the general population were offered to self-test with an AN/MT swab after having been tested by RT-PCR. Here, users were confident that they performed the test correctly and would recommend self-testing to others [30]. Furthermore, interviewing university students and staff when SARS-CoV-2 screening via self-testing was offered at their university [16], C19ST was perceived to reduce the barriers to testing. Nonetheless, some staff and students were also concerned that overoptimistic perceptions of the accuracy of self-testing could potentially lead to change in risk behaviors [39]. Moreover, when implementing self-testing in the routines of a care-home, a significant increase in workload and planning was reported by the care-home staff [34].

## DISCUSSION

Evaluating the clinical utility and epidemiological impact of Ag-RDT based C19ST in 11 studies, we found that C19ST received overall good uptake and may identify additional cases compared to a situation where no dedicated testing is available. In addition, values and preferences indicated that self-testing was generally considered acceptable and feasible, and people perceived it as helpful and easy to perform. However, certainty of evidence was very low for all of the outcomes assessed and the included studies showed a high risk of bias.

Assessing the community level health effect, C19ST achieved a pooled test positivity of 0.2% (95% CI 0.1% to 0.4%, when the alternative was no other dedicated testing in the given setting to be available). Amongst other factors such as the sample type used [9], overall test positivity depends especially on the test accuracy and the pre-test probability of being infected [40]. While most studies deployed Ag-RDTs with similarly high accuracy estimates (Innova and Standard Q; sensitivity 72.2% to 81.4% and specificity 99.1% to 99.3%), for the Ag-RDTs used in the other studies varying accuracy is reported (sensitivity ranging from 39.2% to 77.6% and specificity from 89.2% to 100%) [9]. In addition, the studies’ populations and time during the pandemic (within a surge or not) were very heterogenous, suggesting widely differing pre-test probabilities [18]. Particularly, when pre-test probability is low (between waves), a substantial proportion of positive results may be false positive, even though the Ag-RDT used most frequently in the studies included shows a specificity close to 100% under real-life conditions [41]. For example, at 0.5% prevalence, for a test with 80% sensitivity and 99.5% specificity, only about half of positive tests (44.6%) would be true positive. Thus, the overall proportion of C19ST positives should be interpreted with caution. Nonetheless, given that no other routine testing occurred in these mostly asymptomatic populations, it can be inferred that the cases identified through a positive C19ST result might have not been detected without self-testing or detected only later, when secondary transmission might have already occurred. Therefore, C19ST could have identified cases that might have been missed otherwise, mitigating continued transmission and supporting outbreak control [42,43]. Countries and programs implementing C19ST should focus on using quality-assured products such as those on the WHO emergency list and may also want to consider how to best target C19ST toward those with greater COVID-19 risks and vulnerabilities.

With regards to the individual level health effects, the uptake of C19ST was good overall (53.2% [95% CI 36.7% to 68.9%]), but varied between study populations. The highest uptake (75.7%) was found in health care workers [29]. This is not surprising, as health care workers are likely experienced with COVID-19 testing and their frequent contact with vulnerable populations might foster their motivation to test [44]. In contrast, self-testing uptake was lowest (25.8%) among university students and staff [16]. However, certain operational barriers might have also limited uptake in this study. Students and staff were invited to test only via email and had to download a smartphone app before they could take part in the testing. Self-testing uptake might have been higher if staff and students had been approached in person. Nonetheless, university students and staff might feel less urgence to test for SARS-CoV-2 [45], reducing their motivation compared to health care workers. Further research is needed to investigate where C19ST acceptance is highest, the determinants driving its uptake and possible interventions to support uptake.

In the available studies test results were only applied outside a patient / care-provider interaction. Within the included studies, C19ST was mainly used to inform the need for isolating, potentially offering a much shorter turn-around time to decision than would have been made via rRT-PCR (requiring usually 1 to 2 days until results are communicated). In contrast, a negative self-testing result was mainly followed by the continuation of the status quo. Consequently, self-testing enabled several positive effects outside of health, including people being allowed to leave quarantine for a fixed time interval or continue to work [15,28]. However, there was no evidence to assess the impact of self-testing on virus transmission when used as a tool to release from quarantine [15,28] or when used for mostly asymptomatic screening [15,16,29,34,35]. Especially, when regular C19ST was implemented for care-home staff in parallel to professional testing of visitors with the opening of care-homes to indoor visits, it was not possible to disentangle the impact of C19ST from that of visitor testing [34]. Thus, more studies are needed to evaluate the effect of C19ST when replacing or adding to certain COVID-19 restrictions.

Our study had several strengths. First, it followed rigorous methods, aligning to other WHO commissioned reviews for self-testing, and being supported by an independent methodologist contracted by WHO. In addition, we thoroughly assessed the studies included, utilizing the Newcastle Ottawa Scale with an interpretation guide specific to the review question and following the GRADE approach to evaluate the certainty of evidence. Nonetheless, our study might be limited by its underlying assumptions of C19ST being as accurate as Ag-RDTs when performed by professional users. However, this assumption was based on best current scientific knowledge [13] and is also reflected in the current WHO C19ST guideline [19]. In addition, our study was limited by the small number of studies and data heterogeneity. For several outcomes, performing a meta-analysis and drawing conclusions (e.g., for changes in mortality and morbidity) was not possible. In addition, all studies included were only from high income countries, limiting the representativeness of our results outside of these areas. To foster the representativeness of data, we consider it important for future studies to be done both in high- and low-income countries and across different settings. Moreover, the potential impact of C19ST on the implementation of new test and treat approaches will be important to monitor, as early diagnosis is essential for effective treatment. Also, a better understanding of C19ST’s resource use enhance the comparison between implementing C19ST and professional testing. Finally, studies should adhere to standard guidelines for conducting epidemiological or intervention studies [46,47].

## CONCLUSION

This review found that C19ST can achieve good uptake, and has the potential to identify additional cases that might have gone undiagnosed if no dedicated testing were offered in specific settings outside of health care facilities. C19ST was generally perceived as feasible and easy to use by lay users, but utilizing C19ST to confirm a SARS-CoV-2 infection was not reported. Data was limited for most outcomes and the overall risk of bias was high. The present analysis together with further data presented to the guideline development group led WHO to recommend the use of C19ST as an additional tool to control the COVID-19 pandemic [19]. However, further research, especially in low- and middle-income countries, is needed to thoroughly assess the epidemiological impact of C19ST.

## Supporting information

Newcastle Ottawa Scale assessment

PRISMA checklist

Parameters extracted

Study protocol

Extraction Sheet

Search strategy

Detailed study assessment

Definitions of outcomes assessed

Interpretation guide Newcastle Ottawa Scale

Guide for the GRADE certainty assessment

GRADE certainty assessment

Excluded studies

Outcomes not available

## Data Availability

Data extracted from included studies and used for all analysis is publicly available under https://github.com/stmcg/covid-testing-impact-ma.

## ACKNOWLEDGEMENTS

We highly appreciate the methodological comments and edits from Prof. Elie Akl, Associate Dean for Clinical Research at the American University of Beirut, that strongly improved our study protocol and manuscript. Furthermore, we are very grateful to Prof. Iain Buchan, Chair in Public Health and Clinical Informatics, and Associate Pro Vice Chancellor for Innovation at the University of Liverpool, Dr. David Hughes, Lecturer in Health Data Science at the University of Liverpool, Dr. Paula Parvulescu, Consultant in Public Health Medicine at the Liverpool City Council, Dr. John Tulloch, Tenure Track Fellow at the University of Liverpool, Assoc. Prof. Volker Strenger, Research Lead in Infectiology at the Medical University of Graz, and Prof. Peter Willeit, Professor of Clinical Epidemiology at the Medical University of Innsbruck, for sharing further data from their studies.

## LIST OF ABBREVIATIONS

Ag-RDT: Antigen-based rapid diagnostic tests
C19ST: COVID-19 self-testing
CI: Confidence Interval
GDP: Gross Domestic Product
NOS: Newcastle-Ottawa Scale
rRT-PCR: Real-time reverse transcriptase polymerase chain reaction
WHO: Word Health Organisation

## DECLARATIONS

### Ethics approval and consent to participate

Not applicable.

### Consent for publication

Not applicable.

### Authors’ contributions

LEB: Conceptualization, Data curation, Formal analysis, Investigation, Methodology, Project administration, Resources, Supervision, Validation, Visualization, Writing – original draft, Writing – review & editing

CE: Conceptualization, Data curation, Formal analysis, Investigation, Methodology, Validation, Writing – review & editing

HT: Conceptualization, Data curation, Formal analysis, Investigation, Methodology, Writing – review & editing

SM: Formal analysis, Investigation, Methodology, Validation, Visualization, Writing – review & editing

IDO: Writing – review & editing

SK: Writing – review & editing

SY: Visualization, Writing – review & editing

MG: Software, Writing – review & editing

NRP: Writing – review & editing

BE: Writing – review & editing

AM: Writing – review & editing

SO: Writing – review & editing

JAS: Formal analysis, Investigation, Methodology, Project administration, Writing – original draft, Writing – review & editing

CJ: Formal analysis, Investigation, Methodology, Project administration, Writing – original draft, Writing – review & editing

CD: Conceptualization, Investigation, Methodology, Project administration, Resources, Supervision, Validation, Visualization, Writing – original draft, Writing – review & editing

All authors read and approved the final manuscript.

