## Supplementary material for "The clinical utility and epidemiological impact of self-testing for SARS-CoV-2 using antigen detecting diagnostics: a systematic review and meta-analysis": Newcastle Ottawa Scale assessment

### S1 Fig - Newcastle Ottawa Scale assessment

| Author Journal Year / Dataset | Selection | Comparability | Outcome | Overall |
|---|---|---|---|---|
| Downs JIF 2021 / 1 | ⊕ | | | **1 / 9** |
| Hirst OFID 2021 / 1 | ⊕ ⊕ | | ⊕ ⊕ ⊕ | **5 / 9** |
| Hirst OFID 2021 / 2 | ⊕ ⊕ | | ⊕ ⊕ | **4 / 9** |
| Hoehl Dtsch Arztebl 2021 / 1 | ⊕ | | ⊕ | **2 / 9** |
| Kheiroddin Front. Pediatr. 2021 / 1 | ⊕ | ⊕ | ⊕ ⊕ | **4 / 9** |
| Lamb JIF 2021 / 1 | ⊕ | ⊕ | ⊕ | **3 / 9** |
| Love medRxiv 2021 /1 | ⊕ ⊕ | | ⊕ | **3 / 9** |
| Stohr CMI 2021 / 1 | ⊕ ⊕ | | ⊕ | **3 / 9** |
| Stohr CMI 2021 / 2 | ⊕ ⊕ | | ⊕ | **3 / 9** |
| Tulloch Age Ageing 2021 / 1 | ⊕ | | ⊕ | **2 / 9** |
| University of Liverpool [website] 2021 / 1 | ⊕ | | ⊕ | **2 / 9** |
| University of Liverpool [website] 2021 / 2 | ⊕ ⊕ | | ⊕ ⊕ ⊕ | **5 / 9** |
| Wachinger BMJ peads open 2021 / 1 | ⊕ | | ⊕ | **2 / 9** |
| Wachinger BMJ peads open 2021 / 2 | ⊕ | | ⊕ ⊕ | **3 / 9** |
| Willeit Eurosurveillance 2021 / 1 | ⊕ | | ⊕ ⊕ | **3 / 9** |
| Willeit Eurosurveillance 2021 / 2 | ⊕ | | ⊕ ⊕ | **3 / 9** |
| Willeit Eurosurveillance 2021 / 3 | ⊕ | | ⊕ ⊕ | **3 / 9** |

*Red mark: indicates an overall high risk of bias. Yellow mark: indicates an overall medium risk of bias. Green mark: indicates on overall low risk of bias.*
