## Supplementary material for "The clinical utility and epidemiological impact of self-testing for SARS-CoV-2 using antigen detecting diagnostics: a systematic review and meta-analysis": Study protocol

December 22, 2021

**S1 Text – Study protocol**

**Systematic review on the impact of self-performed Ag-RDTs for SARS-CoV-2 in populations seeking testing**

**BACKGROUND**

With the COVID-19 pandemic continuing to inflict a burden on societies worldwide, self-performed antigen rapid diagnostic tests (Ag-RDTs) for SARS-CoV-2 could be an additional tool to reduce transmission. Self-testing has already been recommended by the World Health Organization (WHO) for various other infectious diseases [1,2]. For SARS-CoV-2, self-testing has a role primarily in the screening of asymptomatic individuals and has been employed for testing to protect (e.g. in high risk settings such as hospitals), to release (e.g. contact testing) and to enable (e.g. regular school or workplace testing) as well as mass-testing (e.g. in Germany). However, the impact has not been systematically evaluated to date. With this systematic review, we aim to assess the impact self-performed Ag-RDTs for SARS-CoV-2 could have on the COVID-19 pandemic.

**PICO QUESTIONS**

What is the impact (O) of implementing self-performed Ag-RDTs for SARS-CoV-2 (I) in populations during the COVID-19 pandemic (P) compared to when no testing or testing with professional use only is performed (C)?

P: persons during the COVID-19 pandemic

I: Self-performed Ag-RDTs for SARS-CoV-2

C: No testing or testing with professional use only performed

O: listed below

**OUTCOMES**

Outcomes to be ranked by GDG and classified into primary and secondary outcomes

(1) Community level health impact
    a. Changes in the transmission of SARS-CoV-2, measured in SARS-CoV-2 incidence, prevalence, cases averted, secondary contacts and secondary cases
    b. Changes in testing uptake / acceptability
(2) Individual level health impact
    a. Changes in COVID-19 morbidity, measured in hospitalization, disease severity
    b. Changes in COVID-19 mortality, measured in deaths averted
    c. Changes in proportion of results reported and test-positivity rate
    d. Changes in time to diagnosis and time to isolate
    e. Changes in proportion taking following action upon receiving test results:
       1) Positive result: linkage with confirmatory testing
       2) Negative result: adherence to public health security measurements (PSHM) and / or seeking testing for other diseases if symptomatic
    f. Changes in any other measurements upon receiving test result
(3) Effects on the health system (hospitalization, ICU occupancy and/or transmission within health care workers)
(4) Effects on not health related aspects: absenteeism, schooling, resource usage, user costs (including opportunity costs) and time-savings
(5) Misuse as use out of the intended or fabrication of data, i.e., reporting false results
(6) Adverse events
(7) Social harms

**INCLUSION CRITERIA**

To be included in the review, an article must meet the following criteria:

1) Study designs that implemented a self-performed Ag-RDT for SARS-CoV-2 in a population seeking testing during the COVID-19 pandemic
2) Measured one or more of the outcomes listed above
3) Study sample size greater 100

No restrictions will be placed based on location of the intervention. We will consider retrospective or prospective cohort or nested cohort studies, case-control or cross-sectional studies, before and after studies, as well as randomized studies.

**VAIRABLES CONSIDERED**

When possible, we will stratify outcomes and present by the following categories.

1) Frequency of testing
   a. Once (without a fixed interval)
   b. Routine, i.e., more than 1 occasion as per a fixed interval
2) SARS-CoV-2 exposure
   a. No known exposure
   b. Known exposure – single
   c. Known exposure – frequent
   d. Unknown / not defined
3) Test distributor: workplace, school/university, publicly available (i.e., “Buergertests)
4) Location of testing: work, home, school/university
5) Costs for persons to be tested: tests free of charge vs. provision required
6) Setting:
   a. Rural or urban,
   b. GDP (categorized by WTO)
   c. literacy of the target population
7) Ag-RDT used: company name, test and lot number (if available), sample type
8) Test assistance: testing was assisted by trained operator, was assisted virtually by trained operator, was assisted in any other way or was not assisted at all
9) Risk level of COVID-19 related morbidity and mortality: as defined by paper
10) Vaccination status: vaccinated vs. unvaccinated
11) Seroconvalescence status: convalescent vs. uninfected
12) Age: persons ≤ 18 years of age, persons 18 – 65 years of age, persons > 65 years of age
13) Misuse, adverse events, social harms: specific to self-testing vs. issues that occur in general for SARS-CoV-2 testing
14) Additional interventions: Any other interventions that were implemented to stop transmission besides self-performed Ag-RDTs for SARS-CoV-2, such as mask wearing, isolation / social distancing practices, travel restrictions

**SEARCH STRATEGY**

The following electronic databases will be searched for any article published between December 1, 2020 and November 30, 2021: PubMed and Web of Science. Secondary reference searching will also be conducted on all studies included in the review. Further, selected experts in the field will be contacted to identify additional articles not identified through other search methods.

The main search terms will be “Severe Acute Respiratory Syndrome Coronavirus 2”, “COVID-19”, “Betacoronavirus”, Coronavirus”, “Self-testing” and “Antigen”. Details on the search algorithm can be found in the appendix. No language restrictions will be applied. Articles in languages other than English will be translated where necessary.

**STUDY SELECTION**

Two reviewers (LEB and HT) will review the titles and abstracts of all publications identified by the search algorithm independently. Afterwards, they will individually conduct a full-text review for those eligible, to select the articles for inclusion in the systematic review. As a final step, they will compare results with each other and a third reviewer (CE). Any disputes will be solved by discussion or by a fourth reviewer (CMD).

**DATA EXTRACTION AND MANAGEMENT**

Data extraction will be performed by one reviewer (HT), using a standardized Google Sheets form, and controlled by a second (LEB). Differences in data extraction will be resolved through consensus or referral to a senior study team member (CMD). Studies that assessed multiple self-performed Ag-RDTs or presented results based on differing parameters (e.g., multiple study locations) will be considered as individual datasets.

The following information will be extracted from each included study:

1) Study identification: Author(s), title, peer-review status, year of publication
2) Study characteristics: type of study, start date of study, end date of study, study location, frequency of testing, test distributor, location of testing, setting, test assistance, rural or urban setting, income, literacy
3) Population characteristics: SARS-CoV-2 exposure, risk factors, vaccination status, history of COVID-19 infection, age, number of females
4) Transmission outcomes: Pre- and post-incidence, -prevalence, -number of hospitalizations, -disease severity, -cases averted, -deaths averted
5) Testing characteristics: Ag-RDT used, sample type, persons tested, number of tests done, number of positive test results, number of invalid test results, test uptake, time to diagnosis, time to isolate, measurements upon receiving test results
6) Other effects: effects on school operations, on quarantine duration, on people’s well-being, absenteeism, resource usage, user costs (including opportunity costs), time-savings and effects on the health system (e.g. ICU bed occupancy)
7) Additional interventions: description of measurements taken besides self-performed Ag-RDTs (both within the study and other general measurements
8) Undesirable effect associated with testing: use out of the intended, fabrication of data, social harm, adverse events

**RISK OF BIAS**

To assess the quality of the included studies, we will employ the Newcastle-Ottawa Scale. The tool consists out of eight questions grouped into three categories (selection, comparability and exposure). Depending on the study’s quality, stars are assigned to each of the questions, leading to an overall zero-star-rating (worst) to nine-star-rating (best) each study [3]. Risk of bias assessment will be performed by two reviewers (LEB and HT) independently.

**GRADING OF EVIDENCE**

Following the GRADE approach [4], if evidence from randomized controlled trials is limited, evidence from non-randomized but controlled studies or observational studies will be used instead, but also downgraded per the GRADE system. Grading of evidence will be performed by two reviewers (LEB and HT) independently and presented via GRADE evidence profiles.

**DATA ANALYSIS**

For community and individual level impact (outcome 1-2), we will prepare forest plots and visually evaluate the heterogeneity between studies. We will assess heterogeneity between studies and assess whether a meta-analysis is feasible. We aim to provide point estimates for changes in incidence, prevalence, number of hospitalizations, disease severity, cases averted and deaths averted along with 95% confidence intervals using a random effects model. Also, for health system and non-health related effects (outcomes 3-4), misuse (outcome 5), adverse events (outcome 6) and social harms (outcome 7) a descriptive analysis will be performed.

**LITERATURE CITED**

1.	World Health Organization. Guidelines on HIV self-testing and partner notification: supplement to consolidated guidelines on HIV testing services. 2016.

2.	World Health Organization. Recommendations and guidance on hepatitis C virus self-testing. 2021.

3.	Wells G, Shea B, O’Connell D, Peterson J, Welch V, Losos M, et al. The Newcastle-Ottawa Scale (NOS) for assessing the quality of nonrandomised studies in meta-analyses. 2013.

4.	Higgins JPT, Thomas J, Chandler J, Cumpston M, Li T, Page MJ, et al. Cochrane Handbook for Systematic Reviews of Interventions version 6.2 (updated February 2021). 2021.

**SEARCH ALGORITHM**

### Topic

Which impact (O) has the implementation of self-performed antigen rapid diagnostics (Ag-RDTs) for SARS-CoV-2 (I) in populations seeking testing (P) compared to when no testing or testing with professional use only (C) is performed?

### Main topic concepts definition

### P

| SARS-CoV-2 |
| --- |

### I

| Selbst-Tests |
| --- |
| antigen |

### Strategy

| 1 | P |
| --- | --- |
| 2 | I |
| 3 | 1 AND 2 |

Publication Date 2020/10/01-2021/11/30

### Databases

- PubMed
- Web of Science Core Collection
- BioRxiv und MedRxiv

### Results report

The results were saved in Endnote and deduplicated. Some articles could still appear more than once.

The hits are sorted by database in Endnote. The PubMed hits were the first to be exported in Endnote. This makes them preferred for deduplication. In other words, in the case of duplicates, entries are removed from other databases.

The number of hits for each database in this report is based on its pre-deduplication status in EndNote.

### PubMed

| **Records number** | **Date** |
| --- | --- |
| 1682 | 13.12.2021 |

### P

| **"Severe Acute Respiratory Syndrome Coronavirus 2"[Supplementary Concept] OR**  **"COVID-19" [Supplementary Concept] OR**  **"Betacoronavirus"[Mesh] OR**  **"Coronavirus"[Mesh] OR**  covid*[tw] OR  coronavirus*[tw] OR  corona virus*[tw] OR  ncov*[tw] OR  "n cov*"[tw] OR  sarscov*[tw] OR  "sars cov*"[tw] OR  2019nCoV*[tw] OR  "2019 nCoV*"[tw] OR  sars2*[tw] OR  "sars 2*"[tw] | 222830 |
| --- | --- |

### I

| **"Self-Testing"[Mesh] OR**  Selftest*[tw] OR  BinaxNow[tw] OR  "Binax Now"[tw] OR  Indicaid[tw] OR  Liaison[tw] OR  Dräger[tw] OR  LumiraDx[tw] OR  Sofia[tw] OR  Hough[tw] OR  Ecotest[tw] OR  Lyher[tw] OR  OnSite[tw] OR  "On Site"[tw] OR  Panbio[tw] OR  RightSign[tw] OR  "Right Sign"[tw] OR  Testsealabs[tw] OR  "V Chek"[tw] OR  CareStart[tw] OR  "Care Start"[tw] OR  iHealth[tw] OR  "i Health"[tw] OR  "BD Veritor"[tw] OR  QuickVue[tw] OR  "Quick Vue"[tw] OR  Ellume[tw] OR  Camtech[tw] OR  Aria[tw] OR  Artron[tw] OR  Fosun[tw] OR  Humasis[tw] OR  Standardiq[tw] OR  "Standard iq"[tw] OR  "Standard i q"[tw] OR  "Green Spring"[tw] OR  Greenspring[tw] OR  Biosynex[tw] OR  "AMP rapid Test"[tw] OR  Innova[tw] OR  Clinitest[tw] OR  GenBody[tw] OR  Genedia[tw] OR  Orawell[tw] OR  Flowflex[tw] OR  Ninonasal[tw] OR  instantsure[tw] OR  "COV S35"[tw] OR  InnoScreen[tw] OR  "My Covid Test"[tw] OR  InteliSwab[tw] OR  DiaTrust[tw] OR  "SG Diagnostics"[tw] OR | 66764 |
| --- | --- |
| ((Home*[tw] OR  self*[tw] OR  personal*[tw] OR  mail*[tw])  AND  Test*[tw]  AND  (**"Antigens"[Mesh] OR**  Antigen*[tw] OR  Lateral flow*[tw] OR  LFA[tw] OR  RDT[tw] OR  rapid*[tw])) |  |

P AND I: 2156

AND (2020/10/1:2021/11/30[pdat]): 1682

### Web of Science Core Collection

| **Records number** | **Date** |
| --- | --- |
| 1040 | 13.12.2021 |

### P

| "covid*" OR  "coronavirus*" OR  "corona virus*" OR  "ncov*" OR  "n cov*" OR  "sarscov*" OR  "sars cov*" OR  "2019nCoV*" OR  "2019 nCoV*" OR  "sars2*" OR  "sars 2*" | 193058 |
| --- | --- |

### I

| Selftest* OR  BinaxNow OR  "Binax Now" OR  Indicaid OR  Liaison OR  Dräger OR  LumiraDx OR  Sofia OR  Hough OR  Ecotest OR  Lyher OR  OnSite OR  "On Site" OR  Panbio OR  RightSign OR  "Right Sign" OR  Testsealabs OR  "V Chek" OR  CareStart OR  "Care Start" OR  iHealth OR  "i Health" OR  "BD Veritor" OR  QuickVue OR  "Quick Vue" OR  Ellume OR  Camtech OR  Aria OR  Artron OR  Fosun OR  Humasis OR  "Standard iq" OR  Standardiq OR  "Standard i q" OR  "Green Spring" OR  Greenspring OR  Biosynex OR  "AMP rapid Test" OR  Innova OR  Clinitest OR  GenBody OR  Genedia OR  Orawell OR  Flowflex OR  Ninonasal  instantsure OR  "COV S35" OR  InnoScreen OR  "My Covid Test" OR  InteliSwab OR  DiaTrust OR  "SG Diagnostics"OR | 81763 |
| --- | --- |
| ((Home* OR  self* OR  personal* OR  mail*)  AND  Test*  AND  (Antigen* OR  "Lateral flow*" OR  LFA OR  RDT OR  rapid*)) |  |

P AND I: 1367

DOP=2020-10-01/2021-11-30: 1040
