## Supplementary material for "The clinical utility and epidemiological impact of self-testing for SARS-CoV-2 using antigen detecting diagnostics: a systematic review and meta-analysis": Definitions of outcomes assessed

**S3 Text – Definitions of outcomes assessed**

| **Outcomes** | **Sub-outcomes** | **Definition** |
| --- | --- | --- |
| (1) Community level health impact | a) Number/proportion of infectious cases detected | Changes in the proportion of positive test. We acknowledge that a positive antigen-based COVID-19 self-testing result does not equal a clinical diagnose by means of real-time reverse transcriptase polymerase chain reaction, and further analyze this in the discussion. |
|  | b) Impact on virus transmission | Changes in the transmission of SARS-CoV-2, measured in SARS-CoV-2 incidence, prevalence, cases averted, secondary contacts and secondary cases |
|  | c) Impact on morbidity and/or mortality | Changes in patient’s morbidity (measured in hospitalization, disease severity) and/or mortality (measured in deaths averted) |
| (2) Individual level health impact | a) Linkage for positive tests | Changes in any actions study participants were required to take after a positive test result |
|  | b) Social harm | Any reduction in an individual’s social well-being |
|  | c) Testing uptake | Changes in the proportion of study participants testing compared to the number of study participants who were offered self-testing and eligible to test |
|  | d) Time to diagnosis | Changes in the time between performing the test and having a final diagnosis on the person’s SARS-CoV-2 status |
|  | e) Testing frequency | Changes in the frequency at which study participants tested |
|  | f) Result reporting | Changes in the proportion of test results reported compared to the overall number of tests performed |
|  | g) Linkage for negatives | Changes in any actions study participants were required to take after a negative test result |
| (3) Broader societal effects: | | Changes in absenteeism, schooling, resource usage, user costs (including opportunity costs) and time-savings |
