## Supplementary material for "The clinical utility and epidemiological impact of self-testing for SARS-CoV-2 using antigen detecting diagnostics: a systematic review and meta-analysis": Interpretation guide Newcastle Ottawa Scale

**S4 Text – Interpretation guide Newcastle Ottawa Scale**

**NEWCASTLE - OTTAWA QUALITY ASSESSMENT SCALE**

**COHORT STUDIES**

Population: persons tested for SARS-CoV-2

Exposure: self-testing for SARS-CoV-2

Non-exposure: no testing or RT-PCR testing regime

Outcome: as listed in the study protocol

Note: A study can be awarded a maximum of one star for each numbered item within the Selection and Outcome categories. A maximum of two stars can be given for Comparability

- Ranking from 0 (lowest) to 9 (highest) start

**Selection**

1) Representativeness of the exposed cohort

a) truly representative (population, random / consecutive enrolment, >80% participation) of the average person tested for SARS-CoV-2 in the community **🟑**

b) somewhat representative (population, random / consecutive enrolment, >80% participation) of the average person tested for SARS-CoV-2 in the community **🟑**

c) selected group of users, e.g., nurses, volunteers

d) no description of the derivation of the cohort

2) Selection of the non-exposed cohort

a) drawn from the same community as the exposed cohort **🟑**

b) drawn from a different source

c) no description of the derivation of the non-exposed cohort or no non-exposed cohort included

3) Ascertainment of exposure

a) secure record (the test results or a picture of such was confirmed by a study official and entered in a
 data record administered by the persons conducting the study) **🟑**

b) somewhat recorded (a picture of the test results was taken by study participants, uploaded into a
 data record administered by the persons conducting the study and controlled by a study official) **🟑**

c) test result was entered into a data record by study participants without any proof or verbally
 communicated to study officials without proof

d) no description

4) Demonstration that outcome of interest was not present at start of study

a) yes **🟑**

b) no

**Comparability**

1) Comparability of cohorts on the basis of the design or analysis

a) study controls for prior infection **🟑**

b) study controls for vaccination status **🟑**

c) did not control

**Outcome**

1) Assessment of outcome

a) epidemiological and/or economic data confirmed by an official authority **🟑**

b) data collected by study officials, e.g., through structured interviews **🟑**

c) self-report

d) no description

2) Was follow-up long enough for outcomes to occur

a) study period was a priori defined **🟑**

b) no

3) Adequacy of follow up of cohorts

a) complete follow up **🟑**

- for routine testing: >80% included participants followed the anticipated testing frequency across the whole study period

- for single-testing: offered testing capacity stayed the same across the whole study period

b) subjects lost to follow up unlikely to introduce bias - small number lost, 60-80% follow up, or
 description provided of those lost **🟑**

c) follow up rate <60% and no description of those lost

d) no statement
