## Supplementary material for "The clinical utility and epidemiological impact of self-testing for SARS-CoV-2 using antigen detecting diagnostics: a systematic review and meta-analysis": Guide for the GRADE certainty assessment

**S5 Text – Guide for the GRADE certainty assessment**

Risk of bias:

- Not serious: included data sets’ median ranking on the NOS was 6 to 9 stars
- Serious: included data sets’ median ranking on the NOS was 3 to 5 stars
- Very serious: included data sets’ median ranking on the NOS was 0 to 2 stars

Indirectness:

We grade indirectness by assessing (1) whether patients may differ from those of interest, (2) whether the intervention differs from the one of interest, (3) whether outcomes of interest are directly measured or through surrogate parameters, and (4) whether the intervention was directly compared to the comparator or this was done relying on a surrogate parameter.

We define patients of interest as average (age, literacy, profession) persons, being a representative sample of low-, middle- and high-income countries.

- Not serious: all data sets directly (i.e., no surrogate parameter had to be used to assess the outcome) evaluated the outcome of interest through the intervention of interest in the study population of interest
- Serious: in at least one data set not all four of the criteria to assess indirectness were met
- Very serious: in a majority of data sets not all four of the criteria to assess indirectness were met

Inconsistency:

We grade inconsistency by evaluating similarity of point estimates, extent of overlap of confidence intervals, and statistical criteria including tests of heterogeneity

- Not serious: all data sets showed similar point estimates, without overlapping confidence interval and low heterogeneity
- Serious: at least one data set showed outstanding point estimates, as also reflected through increased heterogeneity and some confidence intervals were overlapping
- Very serious: point estimates differed widely, as also reflected by high data heterogeneity, and confidence intervals were largely overlapping

Imprecision:

We grade imprecision by evaluating whether using the upper or lower boundary of the outcomes’ confidence intervals would change our recommendation.

- Not serious: in none of the data sets using the upper or lower boundary of the outcomes’ confidence intervals would change our recommendation
- Serious: in at least one of the data sets using the upper or lower boundary of the outcomes’ confidence intervals would change our recommendation
- Very serious: in the majority of data sets using the upper or lower boundary of the outcomes’ confidence intervals would change our recommendation
