## Supplementary material for "The clinical utility and epidemiological impact of self-testing for SARS-CoV-2 using antigen detecting diagnostics: a systematic review and meta-analysis": GRADE certainty assessment

**S6 Text – GRADE assessment**

**Question:** Self-performed Ag-RDT for SARS-CoV-2 compared to no testing or professional testing for populations during the COVID-19 pandemic

The importance of outcomes was decided by the WHO guideline development group. The WHO guideline development group was formed based on nominations to comprise a representative group of diverse expertise, geographic areas and gender. Per WHO policy, all GDG members were assessed for potential conflicts of interest and none were identified. GDG members provided input into the systematic review protocol, including the selection and ranking of outcomes. Using an electronic survey, the GDG rated outcomes on a scale of 1–9 to determine the critical outcomes (1–3: NOT IMPORTANT; 4–6: IMPORTANT; 7–9: CRITICAL). In accordance with the GRADE framework, critical outcomes were the focus the seven highest ranked outcomes were considered in determining the quality of evidence and strength of recommendation. Due to limited evidence, however, all outcomes in evidence profiles.

| **Certainty assessment** | | | | | | | **Impact** | **Certainty** | **Importance** |
| --- | --- | --- | --- | --- | --- | --- | --- | --- | --- |
| **№ of studies** | **Study design** | **Risk of bias** | **Inconsistency** | **Indirectness** | **Imprecision** | **Other considerations** |  |  |  |
| **Individual health outcome - Linkage for positive tests / actions after positive test result** | | | | | | | | | |
| 11^b^ | observational studies | serious^c^ | not serious^d^ | not serious^a^ | not serious^e^ | none | In ten out of the 11 data sets relevant for this outcome, postive results required isolation. In half of these (5 data sets), this was accompanied by the requirement to do an rRT-PCR confirmatory test. In one additional data set, an invitation to conduct an rRT-PCR test was the only measurement reported. | ⨁◯◯◯ Very low | CRITICAL |
| **Individual health outcome - Testing uptake** | | | | | | | | | |
| 5^f^ | observational studies | serious^g^ | not serious^d^ | not serious^a^ | not serious^e^ | none | When only considering data sets where study design offered self-testing as a voluntary option (5 data sets), testing uptake was estimated to be 53.2% (95% CI 36.7% to 68.9%), ranging from 75.7% in hospital staff to 25.8% in university students and staff. | ⨁◯◯◯ Very low | CRITICAL |
| **Individual health outcome - Time to decision** | | | | | | | | | |
| 8^h^ | observational studies | serious^i^ | not serious^d^ | not serious^a^ | not serious^e^ | none | Self-testing usually provides results within 20 to 30 minutes. Self-testing was used to inform the need for isolation in a one-off testing regime (3 data sets) and in regular school testing (5 data sets). In no data sets self-testing was reported to be considered a final SARS-CoV-2 diagnosis, e.g., for clinical decision making or official documentation. | ⨁◯◯◯ Very low | IMPORTANT |
| **Individual health outcome - Result reporting** | | | | | | | | | |
| 17^j^ | observational studies | serious^k^ | not serious^d^ | serious^l^ | not serious^e^ | none | For the majority of data sets (9 data sets), the proportion of results reported was uncertain. In two data sets, study participants were contacted by phone if no test results were submitted, leading to 90.7% results reported. Studies where self-testing was required for study participation (6 data sets; 35.3%), the proportion of results reported was assumed to be 100%. It was not possible to differentiate between the proportion of results reported following positive or negative self-testing results. | ⨁◯◯◯ Very low | IMPORTANT |
| **Community health outcome – Proportion of infectious cases detected / positive test results** | | | | | | | | | |
| 8^m^ | observational studies | serious^n^ | not serious^d^ | not serious^a^ | not serious^e^ | none | The meta-analyzed proportion of positive test results was 0.2% (95% CI 0.1% to 0.4%). Since all eight data sets were conducted in settings where no dedicated testing was performed before, self-testing could have identified cases that might have been missed otherwise. | ⨁◯◯◯ Very low | IMPORTANT |
| **Community health outcome – Impact on virus transmission** | | | | | | | | | |
| 2^o^ | observational studies | serious^p^ | serious^q^ | not serious^a^ | very serious^r^ | none | In one data set it was uncertain whether adding regular self-testing to standard testing and control measures in a care-home impacts the proportion of outbreaks. In the other data set, daily self-testing was used as an alternative to self-quarantine for contacts of cases, but no significant change in secondary cases was detected. | ⨁◯◯◯ Very low | IMPORTANT |
| **Broader societal effects – Impact on absenteeism or economic outputs** | | | | | | | | | |
| 4^s^ | observational studies | serious^t^ | not serious^d^ | not serious^a^ | not serious^e^ | none | In two data sets, where daily self-testing was compared to quarantine and isolation for contacts of confirmed SARS-CoV-2 cases, self-testing reduced the quarantine time to zero, as contacts of confirmed SARS-CoV-2 cases were not required to isolate when providing a negative Ag-RDT self-testing result daily. In one of these data sets, work absenteeism was also reduced, since the persons using self-testing to leave quarantine were police officers, fire fighters, and hospital staff. Self-testing was also reported to increase the wellbeing of staff and children in school. | ⨁◯◯◯ Very low | IMPORTANT |
| **Individual health outcome – Linkage for negatives / actions after nega-tive test result** | | | | | | | | | |
| 10^u^ | observational studies | serious^v^ | not serious^d^ | not serious^a^ | not serious^e^ | none | Negative self-testing results were followed by the continuation of operation (8 data sets) or the permission to leave quarantine (2 data sets). In eigth of these data sets, people had to re-test within a given time interval. | ⨁◯◯◯ Very low | IMPORTANT |

**CI:** confidence interval

#### Explanations

a. Following current guidance from the GRADE guideline, we do not downgrade by one point for all studies but acknowledge that the study populations are not fully representative of the populations of interest. Furthermore, the intervention did not differ from the one of interest and outcomes were reported directly, therefore indirectness was judged 'not serious'.

b. Hirst, J. A., et al., 2021 (2 data sets); Lamb, G., et al., 2021; Love, N., et al., 2021; University of Liverpool, 2021 (2 data sets); Wachinger, J., et al., 2021 (2 data sets); Willeit, P., et al., 2021 (3 data sets)

c. Included data sets showed a comparably high risk of bias on the Newcastle Ottawa Scale (median = 3 stars; Q1 = 3; Q3 = 3.5)

d. For this outcome only qualitative data, or quantitative data in isolated studies in well-described but not comparable settings were available, therefore the criterion 'inconsistency' is negligible and rated as 'not serious'.

e. For this outcome only qualitative data, or quantitative data in isolated studies in well-described but not comparable settings were available, therefore the criterion 'imprecision' is negligible and rated as 'not serious'.

f. Hirst, J.A., et al., 2021; Lamb, G., et al., 2021; Love, N., et al., 2021; Wachinger, J., et al., 2021; Willeit, P., et al., 2021

g. Included data sets show a comparably high risk of bias on the Newcastle Ottawa Scale (median = 3 stars; Q1 = 3; Q3 = 3).

h. Hirst, J.A., et al., 2021 (2 data sets); Lamb, G., et al., 2021; Wachinger, J., et al., 2021 (2 data sets); Willeit, P., et al., 2021 (3 data sets)

i. Included data sets showed a comparably high risk of bias on the Newcastle Ottawa Scale (median = 3 stars; Q1 = 3; Q3 = 3.3)

j. Downs, L.O., et al., 2021; Hirst, J.A., et al., 2021 (2 data sets); Hoehl, S., et al., 2021; Kheiroddin, P., et al., 2021; Lamb, G., et al., 2021; Love, N., et al., 2021; Stohr, J.J.J.M., et al., 2021 (2 data sets); Tulloch, J.S.P., et al., 2021; University of Liverpool, 2021 (2 data sets); Wachinger, J., et al., 2021 (2 data sets); Willeit, P., et al., 2021 (3 data sets)

k. Included data sets showed a comparably high risk of bias on the Newcastle Ottawa Scale (median = 3 stars; Q1 = 2.0; Q3 = 3.0)

l. Following current guidance from the GRADE guideline, we do not downgrade by one point for all studies but acknowledge that the study populations are not fully representative of the populations of interest. Nonetheless, for six data sets, the proportion of results reported had to be estimated based on the fact that self-testing was required for study participants by study design (Kheiroddin, P., et al., 2021; Tulloch, J.S.P., et al., 2021; University of Liverpool, 2021; Willeit, P., et al., 2021 [3 data sets]), therefore indirectness was rated as 'serious'.

m. Hirst, J.A., et al., 2021 (2 data sets); Hoehl, S., et al., 2021; Lamb, G., et al., 2021; 2021; University of Liverpool, 2021 (1 data sets); Willeit, P., et al., 2021 (3 data sets)

n. Included data sets showed a comparably high risk of bias on the Newcastle Ottawa Scale (median = 2.8 stars; Q1 = 3; Q3 = 3.3)

o. Tulloch, J.S.P., et al., 2021 (2 data sets); Love, N., et al., 2021

p. Included data sets showed a comparably high risk of bias on Newcastle Ottawa Scale (median = 2.5 stars; Q1 = 2.3; Q3 = 2.8)

q. Data was limited and heterogeneous, with the two studies reporting different sub-outcomes (proportion of outbreaks: Tulloch, J.S.P., et al., 2021; secondary attack rates: Love, N., et al., 2021). Therefore, inconsistency was ranked as 'serious'.

r. Confidence intervals were too wide (proportion of outbreaks: 54.5% (95% CI 23.4% - 83.3%), Tulloch, J.S.P., et al., 2021; secondary attack rates: 6.3% (95% CI: 3.4% - 11.1%), Love, N., et al., 2021) to precisely judge the effect of the intervention on virus transmission.

s. Love, N., et al., 2021; University of Liverpool, 2021; Wachinger, J., et al., 2021 (2 data sets)

t. Included data sets showed a comparably high risk of bias on the Newcastle Ottawa Scale (median = 2.8 star; Q1 = 3; Q3 = 3.5)

u. Downs, L.O., et al., 2021; Hirst, J.A., et al., 2021 (2 data sets); Love, N., et al., 2021; University of Liverpool, 2021; Wachinger, J., et al., 2021 (2 data sets); Willeit, P., et al., 2021 (3 data sets)

v. Included data sets showed a comparably high risk of bias on the Newcastle Ottawa Scale (median = 3 stars; Q1 = 3; Q3 = 3.8)
