## Supplementary material for "The clinical utility and epidemiological impact of self-testing for SARS-CoV-2 using antigen detecting diagnostics: a systematic review and meta-analysis": Excluded studies

**S7 Text – Excluded studies**

Comment or Review

10.7326/ACPJ202106150-071. Epub 2021 Jun 1.

6.	Soriano V, de Mendoza C, Gómez-Gallego F, Corral O, Barreiro P. Third wave of COVID-19 in Madrid, Spain. Int J Infect Dis, 2021; 107:212-214. DOI:10.1016/j.ijid.2021.04.074

10.1016/j.ijid.2021.04.074. Epub 2021 Apr 27.

7.	Underwood A. COVID-19: A Rural US Emergency Department Perspective. Prehosp Disaster Med, 2021; 36(1):4-5. DOI:10.1017/s1049023x20001417

10.1017/S1049023X20001417. Epub 2020 Nov 4.

Modelling

1.	Chowell G, Dahal S, Bono R, Mizumoto K. Harnessing testing strategies and public health measures to avert COVID-19 outbreaks during ocean cruises. Sci Rep, 2021; 11(1):15482. DOI:10.1038/s41598-021-95032-4

10.1038/s41598-021-95032-4.

2.	Epstein R, Houser C, Wang R. How SARS-CoV-2 and Comparable Pathogens Can Be Defeated in a Single Day: Description and Mathematical Model of the Carrier Separation Plan (CSP). Front Public Health, 2021; 9:640009. DOI:10.3389/fpubh.2021.640009

10.3389/fpubh.2021.640009. eCollection 2021.

3.	Howerton E, Ferrari MJ, Bjørnstad ON, Bogich TL, Borchering RK, Jewell CP, et al. Synergistic interventions to control COVID-19: Mass testing and isolation mitigates reliance on distancing. PLoS Comput Biol, 2021; 17(10):e1009518. DOI:10.1371/journal.pcbi.1009518

10.1371/journal.pcbi.1009518. eCollection 2021 Oct.

4.	Love J, Wimmer MT, Toth DJA, Chandran A, Makhija D, Cooper CK, et al. Comparison of antigen- and RT-PCR-based testing strategies for detection of SARS-CoV-2 in two high-exposure settings. PLoS One, 2021; 16(9):e0253407. DOI:10.1371/journal.pone.0253407

10.1371/journal.pone.0253407. eCollection 2021.

5.	Paltiel AD, Zheng A, Sax PE. Clinical and Economic Effects of Widespread Rapid Testing to Decrease SARS-CoV-2 Transmission. Ann Intern Med, 2021; 174(6):803-810. DOI:10.7326/m21-0510.

6.	Qiu X, Miller JC, MacFadden DR, Hanage WP. Evaluating the contributions of strategies to prevent SARS-CoV-2 transmission in the healthcare setting: a modelling study. BMJ Open, 2021; 11(3):e044644. DOI:10.1136/bmjopen-2020-044644

10.1136/bmjopen-2020-044644.

7.	See I, Paul P, Slayton RB, Steele MK, Stuckey MJ, Duca L, et al. Modeling Effectiveness of Testing Strategies to Prevent Coronavirus Disease 2019 (COVID-19) in Nursing Homes-United States, 2020. Clin Infect Dis, 2021; 73(3):e792-e798. DOI:10.1093/cid/ciab110.

8.	Xie S, Wang W, Wang Q, Wang Y, Zeng D. Evaluating Effectiveness of Public Health Intervention Strategies for Mitigating COVID-19 Pandemic. ArXiv, 2021.

No Ag-RDT

1.	Abdul-Moheeth M, Valencia V, Schaefer S, Brode WM, Nieto K, Moriates C. Improving Healthcare Value: Effectiveness of a Program to Reduce Laboratory Testing for Non-Critically-Ill Patients With COVID-19. J Hosp Med, 2021; 16(8):495-498. DOI:10.12788/jhm.3649

10.12788/jhm.3649.

2.	Abuali M, Bonner R, Irigoyen M. Operationalizing an academic pediatric practice during the COVID-19 crisis. Am J Infect Control, 2021; 49(2):226-228. DOI:10.1016/j.ajic.2020.07.003

10.1016/j.ajic.2020.07.003. Epub 2020 Jul 9.

3.	Bastiani L, Fortunato L, Pieroni S, Bianchi F, Adorni F, Prinelli F, et al. Rapid COVID-19 Screening Based on Self-Reported Symptoms: Psychometric Assessment and Validation of the EPICOVID19 Short Diagnostic Scale. J Med Internet Res, 2021; 23(1):e23897. DOI:10.2196/23897

10.2196/23897.

4.	Borud EK, Nakstad ER, Håberg SE, Lind A, Fadum EA, Taxt AM, et al. Severe acute respiratory syndrome coronavirus 2 prevalence in 1170 asymptomatic Norwegian conscripts. Health Sci Rep, 2021; 4(1):e233. DOI:10.1002/hsr2.233.

5.	Chang JC, Chen YH, Lin MC, Li YJ, Hsu TF, Huang HH, et al. Emergency department response to coronavirus disease 2019 outbreak with a fever screening station and "graded approach" for isolation and testing. J Chin Med Assoc, 2020; 83(11):997-1003. DOI:10.1097/jcma.0000000000000420

10.1097/JCMA.0000000000000420.

6.	Eckardt P, Guran R, Hennemyre J, Arikupurathu R, Poveda J, Miller N, et al. Hospital affiliated long term care facility COVID-19 containment strategy by using prevalence testing and infection control best practices. Am J Infect Control, 2020; 48(12):1552-1555. DOI:10.1016/j.ajic.2020.06.215

10.1016/j.ajic.2020.06.215. Epub 2020 Jul 3.

7.	Escobar DJ, Lanzi M, Saberi P, Love R, Linkin DR, Kelly JJ, et al. Mitigation of a Coronavirus Disease 2019 Outbreak in a Nursing Home Through Serial Testing of Residents and Staff. Clin Infect Dis, 2021; 72(9):e394-e396. DOI:10.1093/cid/ciaa1021

10.1093/cid/ciaa1021.

8.	Fox MD, Bailey DC, Seamon MD, Miranda ML. Response to a COVID-19 Outbreak on a University Campus - Indiana, August 2020. MMWR Morb Mortal Wkly Rep, 2021; 70(4):118-122. DOI:10.15585/mmwr.mm7004a3

10.15585/mmwr.mm7004a3.

9.	Gervasi SF, Pengue L, Damato L, Monti R, Pradella S, Pirronti T, et al. Is extensive cardiopulmonary screening useful in athletes with previous asymptomatic or mild SARS-CoV-2 infection? Br J Sports Med, 2021; 55(1):54-61. DOI:10.1136/bjsports-2020-102789

10.1136/bjsports-2020-102789. Epub 2020 Oct 5.

10.	Hengel B, Causer L, Matthews S, Smith K, Andrewartha K, Badman S, et al. A decentralised point-of-care testing model to address inequities in the COVID-19 response. Lancet Infect Dis, 2021; 21(7):e183-e190. DOI:10.1016/s1473-3099(20)30859-8

10.1016/S1473-3099(20)30859-8. Epub 2020 Dec 24.

11.	Iruzubieta P, Fernández-Lanas T, Rasines L, Cayon L, Álvarez-Cancelo A, Santos-Laso A, et al. Feasibility of large-scale population testing for SARS-CoV-2 detection by self-testing at home. Sci Rep, 2021; 11(1):9819. DOI:10.1038/s41598-021-89236-x.

12.	Liu HH, Ezekowitz MD, Columbo M, Khan O, Martin J, Spahr J, et al. The future is now: our experience starting a remote clinical trial during the beginning of the COVID-19 pandemic. Trials, 2021; 22(1):603. DOI:10.1186/s13063-021-05537-6

10.1186/s13063-021-05537-6.

13.	Loss J, Kuger S, Buchholz U, Lehfeld AS, Varnaccia G, Haas W, et al. [SARS-CoV-2 incidence, transmission, and containment measures in daycare centers during the COVID-19 pandemic-findings from the Corona Daycare Study]. Bundesgesundheitsblatt Gesundheitsforschung Gesundheitsschutz, 2021; 64(12):1581-1591. DOI:10.1007/s00103-021-03449-z.

14.	Mendels DA, Dortet L, Emeraud C, Oueslati S, Girlich D, Ronat JB, et al. Using artificial intelligence to improve COVID-19 rapid diagnostic test result interpretation. Proc Natl Acad Sci U S A, 2021; 118(12). DOI:10.1073/pnas.2019893118

10.1073/pnas.2019893118.

15.	Roberts AT, Wong G, Kotsanas D, Francis MJ, Stuart RL, Graham M, et al. Impact of on-site compared to off-site testing for severe acute respiratory coronavirus virus 2 (SARS-CoV-2) on duration of isolation and resource utilization. Infect Control Hosp Epidemiol, 2021; 42(8):1004-1006. DOI:10.1017/ice.2020.433.

16.	Yokota I, Shane PY, Okada K, Unoki Y, Yang Y, Inao T, et al. Mass Screening of Asymptomatic Persons for Severe Acute Respiratory Syndrome Coronavirus 2 Using Saliva. Clin Infect Dis, 2021; 73(3):e559-e565. DOI:10.1093/cid/ciaa1388.

17.	Zhang Z, Tang Z, Farokhzad N, Chen T, Tao W. Sensitive, Rapid, Low-Cost, and Multiplexed COVID-19 Monitoring by the Wireless Telemedicine Platform. Matter, 2020; 3(6):1818-1820. DOI:10.1016/j.matt.2020.11.001

10.1016/j.matt.2020.11.001.

No relevant outcome

10.1016/j.ijid.2021.08.023. Epub 2021 Aug 18.

8.	Wanat M, Logan M, Hirst JA, Vicary C, Lee JJ, Perera R, et al. Perceptions on undertaking regular asymptomatic self-testing for COVID-19 using lateral flow tests: a qualitative study of university students and staff. BMJ Open, 2021; 11(9):e053850. DOI:10.1136/bmjopen-2021-053850.

9.	Krüger LJ, Klein JAF, Tobian F, Gaeddert M, Lainati F, Klemm S, et al. Evaluation of accuracy, exclusivity, limit-of-detection and ease-of-use of LumiraDx™: An antigen-detecting point-of-care device for SARS-CoV-2. Infection, 2021:1-12. DOI:10.1007/s15010-021-01681-y

10.1007/s15010-021-01681-y.

No self-test

1.	Agulló V, Fernández-González M, Ortiz de la Tabla V, Gonzalo-Jiménez N, García JA, Masiá M, et al. Evaluation of the rapid antigen test Panbio COVID-19 in saliva and nasal swabs in a population-based point-of-care study. J Infect, 2021; 82(5):186-230. DOI:10.1016/j.jinf.2020.12.007.

2.	Alemany A, Baró B, Ouchi D, Rodó P, Ubals M, Corbacho-Monné M, et al. Analytical and clinical performance of the panbio COVID-19 antigen-detecting rapid diagnostic test. J Infect, 2021; 82(5):186-230. DOI:10.1016/j.jinf.2020.12.033

10.1016/j.jinf.2020.12.033. Epub 2021 Jan 7.

3.	Allan-Blitz LT, Klausner JD. A Real-World Comparison of SARS-CoV-2 Rapid Antigen Testing versus PCR Testing in Florida. J Clin Microbiol, 2021; 59(10):e0110721. DOI:10.1128/jcm.01107-21.

4.	Aranaz-Andrés JM, Chávez ACF, Laso AM, Abreu M, Núñez PM, Galán JC, et al. Analysis of the diagnostic accuracy of rapid antigenic tests for detection of SARS-CoV-2 in hospital outbreak situation. Eur J Clin Microbiol Infect Dis, 2021:1-8. DOI:10.1007/s10096-021-04346-8

10.1007/s10096-021-04346-8.

5.	Bachman CM, Grant BD, Anderson CE, Alonzo LF, Garing S, Byrnes SA, et al. Clinical validation of an open-access SARS-COV-2 antigen detection lateral flow assay, compared to commercially available assays. PLoS One, 2021; 16(8):e0256352. DOI:10.1371/journal.pone.0256352

10.1371/journal.pone.0256352. eCollection 2021.

10.1101/2021.01.20.20243782. Preprint.

11.	Chatard JC, Le Gac JM, Gonzalo S, Vaysse P, Coulange M. Management of COVID-19 on board the mixed cargo ship Aranui 5. Int Marit Health, 2021; 72(3):155-162. DOI:10.5603/imh.2021.0031

10.5603/IMH.2021.0031.

12.	Chiu RYT, Kojima N, Mosley GL, Cheng KK, Pereira DY, Brobeck M, et al. Evaluation of the INDICAID COVID-19 Rapid Antigen Test in Symptomatic Populations and Asymptomatic Community Testing. Microbiol Spectr, 2021; 9(1):e0034221. DOI:10.1128/Spectrum.00342-21.

13.	Constantine ST, Callaway D, Driscoll JN, Murphy S. Implementation of Drive-Through Testing for COVID-19 With Community Paramedics. Disaster Med Public Health Prep, 2021:1-7. DOI:10.1017/dmp.2021.46.

14.	Cortés Rubio JA, Costa Zamora MP, Canals Aracil M, Pulgar Feio M, Mata Martínez A, Carrasco Munera A. [Evaluation of the diagnostic test for rapid detection of covid-19 antigen (Panbio Covid rapid test) in primary care]. Semergen, 2021; 47(8):508-514. DOI:10.1016/j.semerg.2021.06.001.

15.	Decker SJ, Goldstein TA, Ford JM, Teng MN, Pugliese RS, Berry GJ, et al. 3-Dimensional Printed Alternative to the Standard Synthetic Flocked Nasopharyngeal Swabs Used for Coronavirus Disease 2019 Testing. Clin Infect Dis, 2021; 73(9):e3027-e3032. DOI:10.1093/cid/ciaa1366

10.1093/cid/ciaa1366.

16.	Delaugerre C, Foissac F, Abdoul H, Masson G, Choupeaux L, Dufour E, et al. Prevention of SARS-CoV-2 transmission during a large, live, indoor gathering (SPRING): a non-inferiority, randomised, controlled trial. Lancet Infect Dis, 2021. DOI:10.1016/s1473-3099(21)00673-3.

17.	Diel R, Nienhaus A. Point-of-care COVID-19 antigen testing in German emergency rooms - a cost-benefit analysis. Pulmonology, 2021. DOI:10.1016/j.pulmoe.2021.06.009

10.1016/j.pulmoe.2021.06.009.

18.	Domínguez Fernández M, Peña Rodríguez MF, Lamelo Alfonsín F, Bou Arévalo G. Experience with Panbio™ rapid antigens test device for the detection of SARS-CoV-2 in nursing homes. Enferm Infecc Microbiol Clin (Engl Ed), 2021. DOI:10.1016/j.eimce.2021.10.002

10.1016/j.eimce.2021.10.002.

19.	Dřevínek P, Hurych J, Kepka Z, Briksi A, Kulich M, Zajac M, et al. The sensitivity of SARS-CoV-2 antigen tests in the view of large-scale testing. Epidemiol Mikrobiol Imunol, 2021; 70(3):156-160.

20.	Ehrenstein B, Schwarz T, Fleck M, Günther F. [Hygiene measures against COVID-19 in routine outpatient care : Acceptance by the patients?]. Z Rheumatol, 2021; 80(4):348-352. DOI:10.1007/s00393-021-00990-9

10.1007/s00393-021-00990-9. Epub 2021 Apr 6.

21.	Eibensteiner F, Ritschl V, Stamm T, Cetin A, Schmitt CP, Ariceta G, et al. Countermeasures against COVID-19: how to navigate medical practice through a nascent, evolving evidence base - a European multicentre mixed methods study. BMJ Open, 2021; 11(2):e043015. DOI:10.1136/bmjopen-2020-043015

10.1136/bmjopen-2020-043015.

22.	Fernández MD, Estévez AS, Alfonsín FL, Arevalo GB. [USEFULNESS OF THE LUMIRADX ™ SARS-COV-2 ANTIGEN TEST IN NURSING HOME]. Enferm Infecc Microbiol Clin (Engl Ed), 2021. DOI:10.1016/j.eimc.2021.06.006

10.1016/j.eimc.2021.06.006.

10.1093/ajcp/aqab081.

10.1016/j.ijid.2021.11.006.

33.	Kanji JN, Proctor DT, Stokes W, Berenger BM, Silvius J, Tipples G, et al. Multicenter Postimplementation Assessment of the Positive Predictive Value of SARS-CoV-2 Antigen-Based Point-of-Care Tests Used for Screening of Asymptomatic Continuing Care Staff. J Clin Microbiol, 2021; 59(11):e0141121. DOI:10.1128/jcm.01411-21.

34.	Kierkegaard P, Micocci M, McLister A, Tulloch JSP, Parvulescu P, Gordon AL, et al. Implementing lateral flow devices in long-term care facilities: experiences from the Liverpool COVID-19 community testing pilot in care homes- a qualitative study. BMC Health Serv Res, 2021; 21(1):1153. DOI:10.1186/s12913-021-07191-9

10.1186/s12913-021-07191-9.

35.	Kim Y, Yu I, Kweon O, Choi JY, Yong D, Park ES. Respiratory Specimen Collection Booth for COVID-19 Test: Efficiency Based Newly Introduced Facility. J Korean Med Sci, 2020; 35(49):e432. DOI:10.3346/jkms.2020.35.e432

10.3346/jkms.2020.35.e432.

36.	Kipritci Z, Keskin A, Çıragil P, Topkaya AE. [Evaluation of a Visually-Read Rapid Antigen Test Kit (SGA V-Chek) for Detection of SARS-CoV-2 Virus]. Mikrobiyol Bul, 2021; 55(3):461-464. DOI:10.5578/mb.20219815

10.5578/mb.20219815.

37.	Kriemler S, Ulyte A, Ammann P, Peralta GP, Berger C, Puhan MA, et al. Surveillance of Acute SARS-CoV-2 Infections in School Children and Point-Prevalence During a Time of High Community Transmission in Switzerland. Front Pediatr, 2021; 9:645577. DOI:10.3389/fped.2021.645577

10.3389/fped.2021.645577. eCollection 2021.

38.	Krüger LJ, Gaeddert M, Tobian F, Lainati F, Gottschalk C, Klein JAF, et al. The Abbott PanBio WHO emergency use listed, rapid, antigen-detecting point-of-care diagnostic test for SARS-CoV-2-Evaluation of the accuracy and ease-of-use. PLoS One, 2021; 16(5):e0247918. DOI:10.1371/journal.pone.0247918

10.1371/journal.pone.0247918. eCollection 2021.

39.	Landaas ET, Storm ML, Tollånes MC, Barlinn R, Kran AB, Bragstad K, et al. Diagnostic performance of a SARS-CoV-2 rapid antigen test in a large, Norwegian cohort. J Clin Virol, 2021; 137:104789. DOI:10.1016/j.jcv.2021.104789.

40.	Mack CD, Osterholm M, Wasserman EB, Petruski-Ivleva N, Anderson DJ, Myers E, et al. Optimizing SARS-CoV-2 Surveillance in the United States: Insights From the National Football League Occupational Health Program. Ann Intern Med, 2021; 174(8):1081-1089. DOI:10.7326/m21-0319.

41.	Mash R, Goliath C, Perez G. Re-organising primary health care to respond to the Coronavirus epidemic in Cape Town, South Africa. Afr J Prim Health Care Fam Med, 2020; 12(1):e1-e4. DOI:10.4102/phcfm.v12i1.2607

10.4102/phcfm.v12i1.2607.

42.	Masiá M, Fernández-González M, Sánchez M, Carvajal M, García JA, Gonzalo-Jiménez N, et al. Nasopharyngeal Panbio COVID-19 Antigen Performed at Point-of-Care Has a High Sensitivity in Symptomatic and Asymptomatic Patients With Higher Risk for Transmission and Older Age. Open Forum Infect Dis, 2021; 8(3):ofab059. DOI:10.1093/ofid/ofab059.

43.	Matsuda EM, de Campos IB, de Oliveira IP, Colpas DR, Carmo A, Brígido LFM. Field evaluation of COVID-19 antigen tests versus RNA based detection: Potential lower sensitivity compensated by immediate results, technical simplicity, and low cost. J Med Virol, 2021; 93(7):4405-4410. DOI:10.1002/jmv.26985

10.1002/jmv.26985. Epub 2021 Apr 8.

44.	McKay SL, Tobolowsky FA, Moritz ED, Hatfield KM, Bhatnagar A, LaVoie SP, et al. Performance Evaluation of Serial SARS-CoV-2 Rapid Antigen Testing During a Nursing Home Outbreak. Ann Intern Med, 2021; 174(7):945-951. DOI:10.7326/m21-0422.

45.	Miller JS, Holshue M, Dostal TKH, Newman LP, Lindquist S. COVID-19 Outbreak Among Farmworkers - Okanogan County, Washington, May-August 2020. MMWR Morb Mortal Wkly Rep, 2021; 70(17):617-621. DOI:10.15585/mmwr.mm7017a3

10.15585/mmwr.mm7017a3.

46.	Moritz ED, McKay SL, Tobolowsky FA, LaVoie SP, Waltenburg MA, Lecy KD, et al. Repeated Antigen Testing Among SARS-CoV-2-Positive Nursing Home Residents. Infect Control Hosp Epidemiol, 2021:1-10. DOI:10.1017/ice.2021.370.

47.	Muhi S, Tayler N, Hoang T, Ballard SA, Graham M, Rojek A, et al. Multi-site assessment of rapid, point-of-care antigen testing for the diagnosis of SARS-CoV-2 infection in a low-prevalence setting: A validation and implementation study. Lancet Reg Health West Pac, 2021; 9:100115. DOI:10.1016/j.lanwpc.2021.100115

10.1016/j.lanwpc.2021.100115. Epub 2021 Mar 2.

48.	Osterman A, Baldauf HM, Eletreby M, Wettengel JM, Afridi SQ, Fuchs T, et al. Evaluation of two rapid antigen tests to detect SARS-CoV-2 in a hospital setting. Med Microbiol Immunol, 2021; 210(1):65-72. DOI:10.1007/s00430-020-00698-8

10.1007/s00430-020-00698-8. Epub 2021 Jan 16.

49.	Patriquin G, Davidson RJ, Hatchette TF, Head BM, Mejia E, Becker MG, et al. Generation of False-Positive SARS-CoV-2 Antigen Results with Testing Conditions outside Manufacturer Recommendations: A Scientific Approach to Pandemic Misinformation. Microbiol Spectr, 2021; 9(2):e0068321. DOI:10.1128/Spectrum.00683-21

10.1128/Spectrum.00683-21. Epub 2021 Oct 20.

50.	Pilarowski G, Lebel P, Sunshine S, Liu J, Crawford E, Marquez C, et al. Performance characteristics of a rapid SARS-CoV-2 antigen detection assay at a public plaza testing site in San Francisco. medRxiv, 2020. DOI:10.1101/2020.11.02.20223891.

51.	Pilarowski G, Marquez C, Rubio L, Peng J, Martinez J, Black D, et al. Field Performance and Public Health Response Using the BinaxNOW (TM) Rapid Severe Acute Respiratory Syndrome Coronavirus 2 (SARS-CoV-2) Antigen Detection Assay During Community-Based Testing. Clinical Infectious Diseases, 2021; 73(9):E3098-E3101. DOI:10.1093/cid/ciaa1890.

52.	Pollock NR, Berlin D, Smole SC, Madoff LC, Brown C, Henderson K, et al. Implementation of SARS-CoV2 Screening in K-12 Schools Using In-School Pooled Molecular Testing and Deconvolution by Rapid Antigen Test. J Clin Microbiol, 2021; 59(9):e0112321. DOI:10.1128/jcm.01123-21

10.1128/JCM.01123-21. Epub 2021 Aug 18.

10.3389/fpubh.2021.708907. eCollection 2021.

58.	Prince-Guerra JL. Evaluation of Abbott BinaxNOW Rapid Antigen Test for SARS-CoV-2 Infection at Two Community-Based Testing Sites (vol 70, pg 103, 2020). Mmwr-Morbidity and Mortality Weekly Report, 2021; 70(4):144-144.

59.	Ralli M, De-Giorgio F, Pimpinelli F, Cedola C, Shkodina N, Morrone A, et al. SARS-CoV-2 infection prevalence in people experiencing homelessness. Eur Rev Med Pharmacol Sci, 2021; 25(20):6425-6430. DOI:10.26355/eurrev_202110_27016

10.26355/eurrev_202110_27016.

60.	Revollo B, Blanco I, Soler P, Toro J, Izquierdo-Useros N, Puig J, et al. Same-day SARS-CoV-2 antigen test screening in an indoor mass-gathering live music event: a randomised controlled trial. Lancet Infect Dis, 2021; 21(10):1365-1372. DOI:10.1016/s1473-3099(21)00268-1.

61.	Routsias JG, Mavrouli M, Tsoplou P, Dioikitopoulou K, Tsakris A. Diagnostic performance of rapid antigen tests (RATs) for SARS-CoV-2 and their efficacy in monitoring the infectiousness of COVID-19 patients. Sci Rep, 2021; 11(1):22863. DOI:10.1038/s41598-021-02197-z

10.1038/s41598-021-02197-z.

10.3389/fpubh.2021.694795. eCollection 2021.

10.1016/j.jhin.2021.03.021. Epub 2021 Mar 27.

72.	van Ogtrop ML, van de Laar TJW, Eggink D, Vanhommerig JW, van der Reijden WA. Comparison of the Performance of the PanBio COVID-19 Antigen Test in SARS-CoV-2 B.1.1.7 (Alpha) Variants versus non-B.1.1.7 Variants. Microbiol Spectr, 2021; 9(3):e0088421. DOI:10.1128/Spectrum.00884-21.

73.	Weigl JAI, Werlang T, Wessendorf M, Helbing H. Vaccine-masked spread of SARS-CoV2 in an elderly care home, and how to prevent a spill-over into the general population. Z Gesundh Wiss, 2021:1-7. DOI:10.1007/s10389-021-01650-7

10.1007/s10389-021-01650-7.

74.	Winkel B, Schram E, Gremmels H, Debast S, Schuurman R, Wensing A, et al. Screening for SARS-CoV-2 infection in asymptomatic individuals using the Panbio COVID-19 antigen rapid test (Abbott) compared with RT-PCR: a prospective cohort study. BMJ Open, 2021; 11(10):e048206. DOI:10.1136/bmjopen-2020-048206.

75.	Xu J, Suo W, Goulev Y, Sun L, Kerr L, Paulsson J, et al. Handheld Microfluidic Filtration Platform Enables Rapid, Low-Cost, and Robust Self-Testing of SARS-CoV-2 Virus. Small, 2021:e2104009. DOI:10.1002/smll.202104009.

76.	Zellmer S, Ebigbo A, Kahn M, Muzalyova A, Classen J, Grünherz V, et al. Evaluation of the ESGE recommendations for COVID-19 pre-endoscopy risk-stratification in a high-volume center in Germany. Endosc Int Open, 2021; 9(10):E1556-e1560. DOI:10.1055/a-1526-1169

10.1055/a-1526-1169. eCollection 2021 Oct.

Study sample size smaller / equal 100

1.	Denford S, Martin AF, Love N, Ready D, Oliver I, Amlôt R, et al. Engagement With Daily Testing Instead of Self-Isolating in Contacts of Confirmed Cases of SARS-CoV-2: A Qualitative Analysis. Front Public Health, 2021; 9:714041. DOI:10.3389/fpubh.2021.714041

10.3389/fpubh.2021.714041. eCollection 2021.

2.	Molnár D, Zsigmond F, Helfferich F. Safety Precautions for Self-Performed Severe Acute Respiratory Syndrome Coronavirus 2 Tests: A Case of a Swallowed Swab. Cureus, 2021; 13(5):e15297. DOI:10.7759/cureus.15297

10.7759/cureus.15297.

3.	Wyman MT, Symms J, Viscusi C. Nasal Foreign Body, an Unanticipated Complication of COVID-19 Care: A Case Report. J Emerg Med, 2021; 60(6):e141-e145. DOI:10.1016/j.jemermed.2020.12.034

10.1016/j.jemermed.2020.12.034. Epub 2020 Dec 28.

4.	Frediani JK, Levy JM, Rao A, Bassit L, Figueroa J, Vos MB, et al. Multidisciplinary assessment of the Abbott BinaxNOW SARS-CoV-2 point-of-care antigen test in the context of emerging viral variants and self-administration. Sci Rep, 2021; 11(1):14604. DOI:10.1038/s41598-021-94055-1.

5.	Nikolai O, Rohardt C, Tobian F, Junge A, Corman VM, Jones TC, et al. Anterior nasal versus nasal mid-turbinate sampling for a SARS-CoV-2 antigen-detecting rapid test: does localisation or professional collection matter? Infect Dis (Lond), 2021; 53(12):947-952. DOI:10.1080/23744235.2021.1969426.

Protocol

1.	Cozzolino I, Ronchi A, Franco R. Rapid on-site evaluation and the COVID-19 pandemic. Cancer Cytopathol, 2020; 128(12):910. DOI:10.1002/cncy.22297.

2.	Setiabudi W, Hungerford D, Subramaniam K, Vaselli NM, Shaw VE, Wilton M, et al. Prospective observational study of SARS-CoV-2 infection, transmission and immunity in a cohort of households in Liverpool City Region, UK (COVID-LIV): a study protocol. BMJ Open, 2021; 11(3):e048317. DOI:10.1136/bmjopen-2020-048317.

3.	Shilton S, Ivanova Reipold E, Roca Álvarez A, Martínez-Pérez GZ. Assessing Values and Preferences Toward SARS-CoV-2 Self-testing Among the General Population and Their Representatives, Health Care Personnel, and Decision-Makers: Protocol for a Multicountry Mixed Methods Study. JMIR Res Protoc, 2021; 10(11):e33088. DOI:10.2196/33088

10.2196/33088.

4.	Bien-Gund CH, Shah J, Ho JI, Stephens-Shields A, Shea K, Fishman J, et al. The COVID-19 Self-Testing through Rapid Network Distribution (C-STRAND) trial: A randomized controlled trial to increase COVID-19 testing in underserved populations. Contemp Clin Trials, 2021; 110:106585. DOI:10.1016/j.cct.2021.106585.

Survey without test impelementation

1.	Betsch C, Sprengholz P, Siegers R, Eitze S, Korn L, Goldhahn L, et al. Empirical evidence to understand the human factor for effective rapid testing against SARS-CoV-2. Proc Natl Acad Sci U S A, 2021; 118(32). DOI:10.1073/pnas.2107179118

10.1073/pnas.2107179118.

2.	Bien-Gund C, Dugosh K, Acri T, Brady K, Thirumurthy H, Fishman J, et al. Factors Associated With US Public Motivation to Use and Distribute COVID-19 Self-tests. JAMA Netw Open, 2021; 4(1):e2034001. DOI:10.1001/jamanetworkopen.2020.34001.

3.	Blake H, Knight H, Jia R, Corner J, Morling JR, Denning C, et al. Students' Views towards Sars-Cov-2 Mass Asymptomatic Testing, Social Distancing and Self-Isolation in a University Setting during the COVID-19 Pandemic: A Qualitative Study. Int J Environ Res Public Health, 2021; 18(8). DOI:10.3390/ijerph18084182

10.3390/ijerph18084182.

4.	Boulliat C, Bilong CV, Dussart C, Massoubre B. [Use of self-tests and rapid diagnostic tests: Survey of dispensing pharmacists in the Auvergne-Rhône-Alpes region]. Ann Pharm Fr, 2021; 79(5):547-557. DOI:10.1016/j.pharma.2021.01.011.

5.	Clipman SJ, Wesolowski AP, Gibson DG, Agarwal S, Lambrou AS, Kirk GD, et al. Rapid Real-time Tracking of Nonpharmaceutical Interventions and Their Association With Severe Acute Respiratory Syndrome Coronavirus 2 (SARS-CoV-2) Positivity: The Coronavirus Disease 2019 (COVID-19) Pandemic Pulse Study. Clin Infect Dis, 2021; 73(7):e1822-e1829. DOI:10.1093/cid/ciaa1313.

6.	Dreyer NA, Reynolds M, DeFilippo Mack C, Brinkley E, Petruski-Ivleva N, Hawaldar K, et al. Self-reported symptoms from exposure to Covid-19 provide support to clinical diagnosis, triage and prognosis: An exploratory analysis. Travel Med Infect Dis, 2020; 38:101909. DOI:10.1016/j.tmaid.2020.101909

10.1016/j.tmaid.2020.101909. Epub 2020 Nov 3.

7.	Goggolidou P, Hodges-Mameletzis I, Purewal S, Karakoula A, Warr T. Self-Testing as an Invaluable Tool in Fighting the COVID-19 Pandemic. J Prim Care Community Health, 2021; 12:21501327211047782. DOI:10.1177/21501327211047782.

8.	Graziadio S, Urwin SG, Cocco P, Micocci M, Winter A, Yang Y, et al. Unmet clinical needs for COVID-19 tests in UK health and social care settings. PLoS One, 2020; 15(11):e0242125. DOI:10.1371/journal.pone.0242125

10.1371/journal.pone.0242125. eCollection 2020.

9.	Haque M, Ferdous AS, Miller J, Linke JA, Dixon C, Athan E, et al. Barriers to performing onsite COVID-19 testing during the second wave in Victoria: Experiences of general practices. Aust J Gen Pract, 2021; 50(11):845-849. DOI:10.31128/ajgp-05-21-6003

10.31128/AJGP-05-21-6003.

10.	Lin L, Song Y, Wang Q, Pu J, Sun FY, Zhang Y, et al. Public Attitudes and Factors of COVID-19 Testing Hesitancy in the United Kingdom and China: Comparative Infodemiology Study. JMIR Infodemiology, 2021; 1(1):e26895. DOI:10.2196/26895

10.2196/26895. eCollection 2021 Jan-Dec.

11.	Massaccesi C, Chiappini E, Paracampo R, Korb S. Large Gatherings? No, Thank You. Devaluation of Crowded Social Scenes During the COVID-19 Pandemic. Front Psychol, 2021; 12:689162. DOI:10.3389/fpsyg.2021.689162

10.3389/fpsyg.2021.689162. eCollection 2021.

12.	Mouliou DS, Pantazopoulos I, Gourgoulianis KI. Societal Criticism towards COVID-19: Assessing the Theory of Self-Diagnosis Contrasted to Medical Diagnosis. Diagnostics (Basel), 2021; 11(10). DOI:10.3390/diagnostics11101777.
